# Neurobehavioral Assessment of Sensorimotor Function in Autism Using Smartphone Technology

**DOI:** 10.1101/2025.07.25.25331276

**Authors:** Kayleigh D. Gultig, Cornelis P. Boele, Lotte E.M. Roggeveen, Ting Fang Soong, Seth Sherry, Caroline Jung, Sara Milosevska, Anton Uvarov, Khalid Benhassan, Said Ait BenAli, Valeria Carpio-Arias, Sander Lindemann, Sebastiaan K.E. Koekkoek, Esra Sefik, Myrthe J. Ottenhoff, Samuel S.-H. Wang, Chris I. De Zeeuw, Abdeslem El Idrissi, Henk-Jan Boele

## Abstract

Differences in sensorimotor processing represent an important, yet underrecognized, feature of autism; typically assessed through subjective observations, which are susceptible to biases.

A more objective approach to quantify sensorimotor function may be possible through reflex-based neurobehavioral evaluations. The clinical application of these assessments has, however, been confined largely to laboratory settings. Thus, small sample sizes and inconsistent findings have made it challenging to understand how sensorimotor function differs in autism and whether it can be used as an objective biomarker for diagnostics.

Here we present a novel smartphone-based platform to conduct neurobehavioral evaluations by measuring facial and behavioral responses in at-home environments. Through a multi-centre study, we explored the platform’s ability to distinguish between children with and without autism.

We enrolled 536 children aged 3–12 years. BlinkLab smartphone-based assessments were successfully completed in 431 children (80.4%), including 275 with autism and 156 neurotypical children. We found that autistic children showed altered sensorimotor responses across multiple domains. These included reduced prepulse inhibition (PPI), stronger habituation over the course of a PPI test, more variable eyeblink responses to auditory stimuli and significant sensitization. Additionally, children with autism displayed more screen avoidance, postural instability, head movements, mouth openings, non-syllabic vocalizations, horizontal pupil shifts, "side-eyeing", and variation in baseline eyelid opening. Exploratory analyses showed that these effects were largely independent of co-occurring ADHD or intellectual disability. Notably, co-occurrence did influence certain subdomains (e.g., PPI, mouth openings).

These findings illustrate that smartphone-based assessments can capture distinct sensorimotor profiles associated with autism in real-world environments.

## Introduction

Atypical sensorimotor functioning is highly prevalent in autism, however, limited attention has been given to sensorimotor skills in both autism research and clinical practice (Coll et al., 2020). Importantly, sensorimotor skills are strongly associated with the level of functioning in autism (Hannant et al., 2016; Patterson et al., 2022; Travers et al., 2017) and more specifically with cognitive abilities (Denisova & Wolpert, 2024). Advancements in digital technology have made the quantification of such sensorimotor skills possible through techniques such as computer vision analysis. These quantifications could complement existing established tools in the autism diagnostic procedure including structured interviews and questionnaires. Objective biomarkers for autism could potentially contribute to more efficient autism diagnostic pathways (Kanne & Bishop, 2021).

An interesting potential biomarker that has not yet been studied with computer vision analysis includes neurobehavioral evaluations assessing sensorimotor function that rely on brain reflexes. Examples include the acoustically evoked eyelid startle reflex (ASR) which can be modified in prepulse inhibition (PPI) to measure sensorimotor gating (Braff et al., 2001; Swerdlow et al., 1999). Here, the response to a normally startling stimulus is reduced by a preceding weaker stimulus (Braff et al., 2001; Swerdlow et al., 1999). Another example, is startle habituation (HAB), a decreased response in startle amplitude with repeated stimulus presentations (Pilz & Schnitzler, 1996), which represents a basic form of non-associative learning (López-Schier, 2019; Simons-Weidenmaier et al., 2006). PPI and HAB can be used to understand different aspects of sensory information processing (Abel et al., 1998; Braff et al., 2001; Graham, 1975; Koch, 1999; Madsen et al., 2014; Perry et al., 2007; Swerdlow et al., 1999). Differences in PPI and HAB between autism and neurotypical development are found in pre-clinical autism research in animal models (El-Cheikh Mohamad et al., 2023) as well as human studies (Cheng et al., 2018; Kohl et al., 2014; Madsen et al., 2014; McAlonan et al., 2002; Perry et al., 2007); however, these findings are not consistent (Cheng et al., 2018; Oranje et al., 2013; Ornitz et al., 1993; Takahashi et al., 2016, 2017; Yuhas et al., 2011), likely due to small sample sizes, variability in stimulus intensities and participant characteristics (Cheng et al., 2018; Doornaert et al., 2024).

Another type of biomarker could be more general sensorimotor behaviors also observable in neurobehavioral assessments and easily quantifiable with computer vision analysis. Early identification of autism is important for optimal clinical outcomes (Franz et al., 2022; Zwaigenbaum et al., 2015) and while differences in motor development are not described as a core feature of autism in the DSM-V, they are reported to be visible in earlier stages of development (Patterson et al., 2022; West, 2019; Wilson et al., 2021) compared to social and language development. Differences in motor and sensorimotor behavior are supported by a substantial body of recent evidence. For example, differences in head-and-body movements (Campbell et al., 2019; Dawson et al., 2018; Krishnappa Babu et al., 2023; Martin et al., 2018; Zhao et al., 2021, 2022) as well as vocalizations (Tenenbaum et al., 2020) have been shown in children with autism compared to neurotypical children.

Until recently, the scalability and potential clinical utility of detailed neurobehavioral assessments to measure these sensorimotor reflexes and behaviors have been limited by the need for specialized lab-bound equipment (Cheng et al., 2018; Dwyer et al., 2023; Madsen et al., 2014). We have developed a user-friendly smartphone-based platform specifically designed for conducting neurobehavioral evaluations, called BlinkLab (**figure 1**). BlinkLab is optimized for at-home use, yielding robust results across various neurobehavioral tests and providing the opportunity to quantify sensorimotor functioning in large cohorts (Boele et al., 2023).

**Figure 1.**
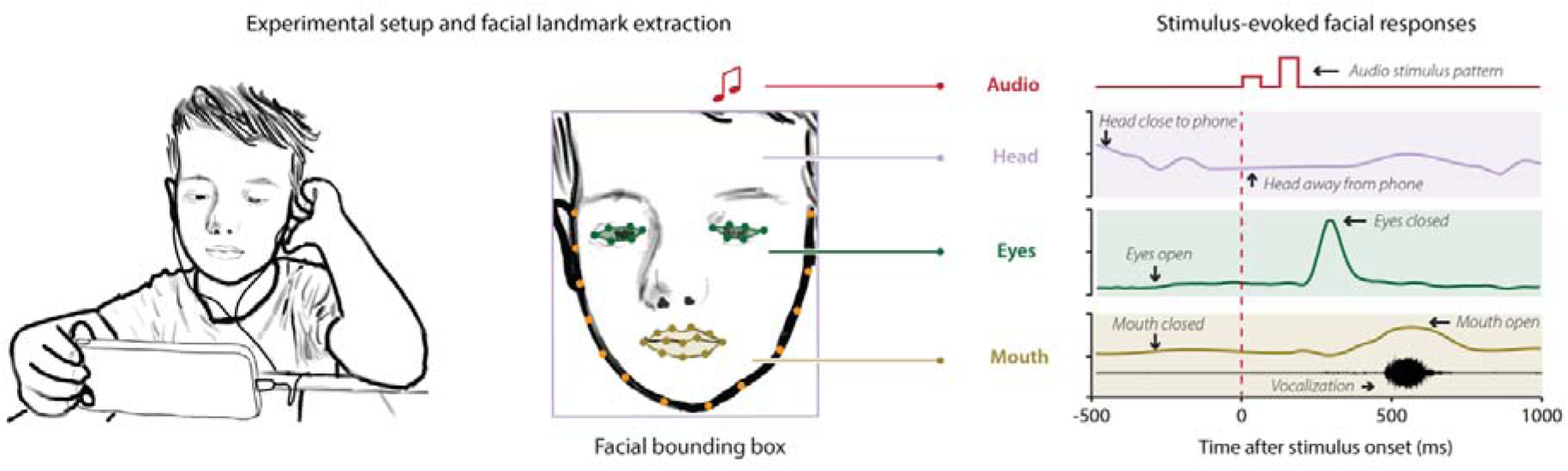
| Data were collected using a smartphone-based platform for neurobehavioral evaluations Children watched a 15-minute video on a smartphone during which brief auditory stimulus patterns were delivered. The smartphone’s camera captured the child’s postural, head, facial, and vocal responses before and after stimulus onset. Illustrations represent a subset of tracked responses.

Here, we performed a retrospective case-control study to test whether BlinkLab smartphone-based neurobehavioral assessments of sensorimotor function showed differences between children with autism and neurotypical children. We hypothesized that neurobehavioral measures including the ASR, PPI, and HAB, would differ in autistic compared to neurotypical children. While analyzing these reflex-based measures, we noticed distinct behaviors that more frequently occurred in children with autism compared to neurotypical children. Given the substantial body of evidence on differences in motor and sensorimotor behaviors in autism, we used our observations to guide our second, data-driven, aim where we compared sensorimotor behaviors in autistic and neurotypical children.

## Methods and Materials

### Participants

A cohort of 536 participants aged between 3 and 12 years was recruited (table 1). Children diagnosed with autism, comprising 114 girls and 256 boys, were recruited at the Mohammed VI National Center for the Disabled (CNMH) at all eight locations in Morocco, including Fes, Salé, Safi, Marrakesh, Casablanca, Oujda, Tangier, and Agadir. These locations provided us with the opportunity to test unmedicated children in a population that is underrepresented in autism research (Durkin et al., 2015). The autism diagnosis was established prior to recruitment by a multidisciplinary team of specialists. The diagnostic procedure started with clinical observations based on symptoms observed and parent reports. Children were then referred to a pediatrician or neurologist to rule out any medical causes. Diagnoses are made according to the Diagnostic and Statistical Manual of Mental Disorders (American Psychiatric Association, D. S. M. T. F., & American Psychiatric Association, 2013) criteria with different scales such as the Vineland Adaptive Behavior Scale used to evaluate different behavioral domains. Neurotypical controls, consisting of 72 girls and 94 boys without a neuropsychiatric diagnosis, were recruited from two schools, one in Taounate (90 km from Fes) and the other in Salé. All participants were selected regardless of sex, gender identity or race. Excluded were participants using medication that affects the nervous system (classified as ATC N0 medication, https://www.whocc.no), as such medication may modulate the acoustically evoked eyelid startle reflex (Geyer et al., 2001; Kumari et al., 2001). Written informed consent was obtained from the parents or caregivers of the children involved in the study, and verbal assent was given by the participating children, except in cases where children were non-verbal. The study was performed in accordance with relevant guidelines and regulations and was reviewed and approved by the institutional review boards of Princeton University (#13943) and the Faculté de Médecine et de Pharmacie de Marrakech in Morocco (#23/2022).

**Table 1.** | Demographic and clinical characteristics of participants

|  | All participants | Successful |  | Rejected |  |
| --- | --- | --- | --- | --- | --- |
|  |  | Autism | Neurotypical | Autism | Neurotypical |
| **Variable** | N = 536 | N = 275 (51.31%) | N = 156 (29.10%) | N = 95 (17.72%) | N = 10 (1.87%) |
| Girls | 186 (34.70%) | 79 (28.73%) | 69 (44.23%) | 35 (36.84%) | 3 (30%) |
| Boys | 350 (65.30%) | 196 (71.27%) | 87 (55.77%) | 60 (63.16%) | 7 (70%) |
| **Age in years (mean ± SD)** | 7.51 (± 2.10 SD) | 8.05 (± 1.89 SD) | 6.97 (± 2.11 SD) | 6.94 (± 2.14 SD) | 6.20 (± 2.61 SD) |
| **Age range (years)** |  |  |  |  |  |
| 3-5 | 144 (26.87%) | 47 (17.09%) | 60 (38.46%) | 32 (33.68%) | 5 (50.00%) |
| 6-8 | 241 (44.96%) | 133 (48.36%) | 68 (43.59%) | 37 (38.95%) | 3 (30.00%) |
| 9-12 | 151 (28.17%) | 95 (34.55%) | 28 (17.95%) | 26 (27.37%) | 2 (20.00%) |
| **Reason for rejection** |  |  |  |  |  |
| Volume | - | - | - | 15 (15.79%) | 1 (10.00%) |
| Headphone refusal/agitation | - | - | - | 35 (36.84%) | 3 (30.00%) |
| Premature test termination | - | - | - | 45 (47.37%) | 6 (60.00%) |

### Experimental setup

Neurobehavioral testing was performed using BlinkLab, a smartphone-based platform (Boele et al., 2023). The tests included specific neurometric tests, including the ASR, PPI and long-term HAB (Test I), and short-term HAB (Test II) along with general measurements of spontaneous and stimulus-evoked postural, head, facial, and vocal responses (Tests I and II) (Abel et al., 1998; Dawson et al., 2018; Graham, 1975; Krishnappa Babu et al., 2023; Martin et al., 2018; Swerdlow et al., 1999; Tenenbaum et al., 2020) . Specifically, Test I consisted of pulse-only trials as well as prepulse (of varying intensities from 5 to 25% of the intensity of the pulse) + pulse trials described in more detail in the supplementary methods. Test two consisted of two habituation paradigms namely a rhythmic (constant interval between stimuli) and random (inconsistent interval between stimuli) delivery of six white noise pulses (**supplementary fig. 1**). Children participated in two 15-minute tests **(figure 1, supplementary fig. 2)**. During each test, the children watched an audio-normalized movie, meaning the volume of the video’s audio track was adjusted to ensure its peak level did not exceed a 60-70 dB sound pressure level, while the trials containing the auditory stimuli were delivered via headphones. Audio normalization ensured that the audio remained within a desired dynamic range, preventing distortion or clipping and maintaining a consistent volume level, serving as ‘background’ against the audio white noise bursts that were delivered during the smartphone neurobehavioral tests. For each trial, computer vision algorithms were used to track and record the position of the participant’s facial landmarks over time.

### Outcome measures

For the analysis of the neurometric data, we used a similar data analysis pipeline to Boele and colleagues (Boele et al., 2023), described in detail in the supplementary methods. The following outcome measures were analyzed from Test I: the amplitude of the spontaneous blink normalized eyelid closure (NEC) response to pulse-only trials (ASR), and prepulse + pulse trials (PPI), as well as amplitude of the ASR over the course of the PPI experiment (long-term HAB). From Test II we analysed the amplitude of NEC in response to the first pulse compared to the next five pulses for both the rhythmic and random stimulus patterns. We also analyzed the anticipatory eye blinks (AEB) in predefined observation windows (OW) containing an omitted pulse (**supplementary fig. 1**). AEB were responses that were not triggered by the startle stimulus itself but rather by the expectation of an upcoming startle stimulus. The cumulative sum of the NEC amplitude was also determined for the rhythmic and random HAB patterns. For all reflexive and anticipatory blinks, we calculated the subject median averaged eyelid closure (NEC) amplitude and variability (standard deviation).

Computer vision algorithms tracking and recording the position of the participants’ facial landmarks over time allowed us to perform a data-driven behavioral analysis where we began by listing behaviors that occurred more frequently in children with autism compared to neurotypical children. These behaviors included: general body movements, moving out of the phone camera view, head touches, headphone touches, pupil movements, mouth opening and vocalizations. We refined these behaviors to be clearly distinguishable and easily quantifiable. This resulted in 10 potential biomarkers, described in more detail in the supplementary methods section, where we quantified: 1. The percentage of trials wherein the child was not detectable in the smartphone’s screen and camera. 2. The percentage of trials wherein the child was touching the headphones. 3. The percentage of trials wherein the child produced non-syllabic vocalizations and the corresponding duration of occurrence. 4. The child’s anteroposterior postural stability. 5. The percentage of trials wherein the child was rotating their head. 6. The size and duration of the child’s mouth openings and closings. 7. The mean horizontal pupil movement per session. 8. How much a participant’s pupil movement was in the opposite direction from the direction their head was turned. 9. The variation in baseline (pre-stimulus) eyelid movements in the first 5 trials of a session. 10. The stability of the variation in baseline eyelid movements across a session.

### Statistical analysis

To test for age differences between neurotypical and autistic children, we used the Wilcoxon rank sum test with continuity correction. Sex differences between the two diagnostic groups were tested using a chi-square test. An ANOVA was used to test for age differences between children with autism with and without co-occurring conditions.

Multilevel linear mixed effects (LME) models were used to assess the following group differences: NEC amplitude for the different PPI stimulus types, amplitude of AEBs in HAB experiments, and the maximum cumulative sum of NEC amplitude in a HAB experiment. ANOVA on LME models were run for within group analyses of the effect of stimulus type on NEC amplitude (PPI), the effect of trial number on NEC amplitude (long-term HAB) between and within groups, the effect of pulse number and group on NEC amplitude (short-term HAB). Linear regression models were used to assess group differences in NEC amplitude variability for the different PPI stimulus types, differences in Pearson correlation coefficients for regression of the ASR during PPI and differences in the variability of the maximum cumulative sum of NEC amplitude in a HAB experiment.

Multilevel binomial logistic regression was used to compare group differences in the trial-level occurrence of the following behaviors: screen avoidance, headphone touches and non-syllabic vocalizations. For all other behaviors, linear regression models were used.

For more experimental and statistical details, we refer to the **Supplementary Methods.**

## Results

We enrolled a total of 536 children between the ages of 3 and 12 years old, spanning the period between May 15, 2023, and December 7th, 2024. BlinkLab tests were successfully administered in 431 (80.41%) children of which 275 (63.81%) had autism and 156 (36.19%) were neurotypical (table 1). Reasons for rejection are listed in table 1. The children with autism were significantly older than the neurotypical children (W = 14472, p < 0.0001, r = - 0.27, table 1) and there was a significant difference in the proportion of males and females between the two groups (χ*2* (1) = 10.61, p = 0.0011). To control for any possible effects of age or sex we conducted all analyses with and without age and sex as a confounder in all models and reported where age or sex had a significant effect.

### Acoustic startle response (ASR)

We did not detect a significant difference in the amplitude of the NEC amplitude for pulse-only trials between autistic and neurotypical children (b = 0.05, t_381_=1.82, CI [-0.004 - 0.09], p=0.070; **figure 2A** **supplementary table 1A)**. The mean eyelid startle amplitude for children with autism was 0.33 (±0.34), while in neurotypical children it was 0.29 (±0.31). Sex was a significant confounder (b = -0.06, t_380_=-2.52, CI [- 0.11 - -0.01] p=0.012; **supplementary table 1A**), however, age was not. In the model including sex, diagnosis did have a significant effect on the amplitude of NEC (b = 0.05, t_380_= 2.19, CI [0.01 - 0.10], p = 0.029).

**Figure 2.**
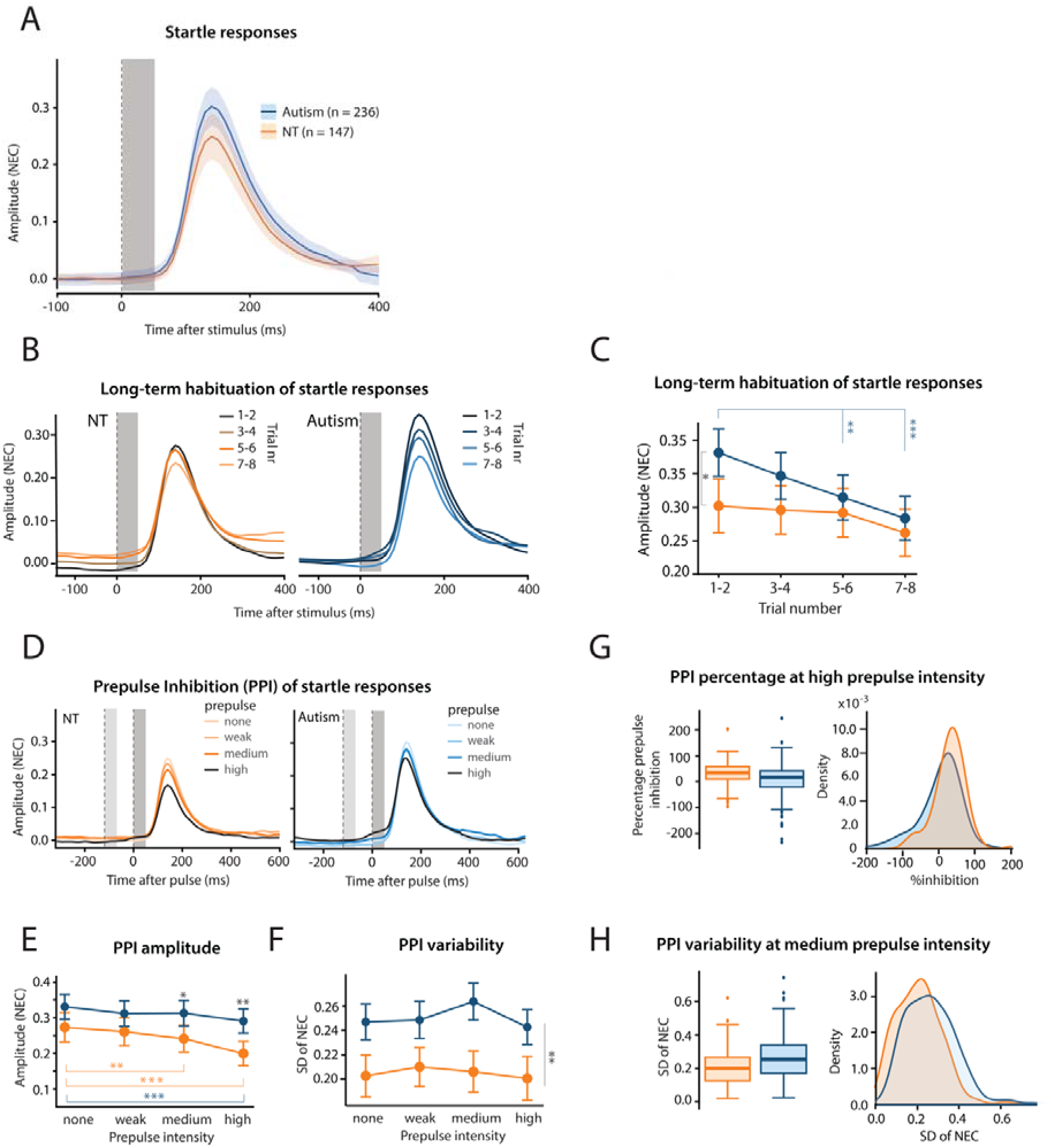
| Characteristics of the auditory startle response, long-term startle habituation, and prepulse inhibition in children with autism and neurotypical (NT) children. **(A)** Group averaged eyelid movement plots showing the acoustically evoked startle responses for children with autism (blue) and NT children (orange). At a group level, children with autism and NT children had comparable eyelid startle responses, however, when sex was accounted for, children with autism had significantly higher startle responses than NT children. **(B)** Group averaged eyelid movement plots of the pulse-only trials ordered by trial number. Trials 1-2 (dark coloring) were delivered at the start and trials 7-8 (light coloring) at the end of the 15-minute PPI smartphone test. **(C)** Mean eyelid startle amplitude as a function of trial number for children with autism and NT children. Children with autism showed significant long-term HAB whereas NT children did not. **(D)** Group averaged eyelid movement plots ordered by stimulus type, i.e. by the intensity of the prepulse that preceded the pulse. For panels A-B and E, the left vertical dashed line indicates the onset of the auditory prepulse, the right one indicates the onset of the pulse. Gray vertical columns indicate the stimulus durations of 50 ms. **(E)** Mean amplitude of the eyelid startle response as a function of prepulse intensity. The group with NT children exhibited a typical prepulse inhibition (PPI) response, characterized by a decrease in mean eyelid startle amplitude as the intensity of the prepulse increased. The group of children diagnosed with autism showed some evidence of PPI, however, only for the highest prepulse intensity trials. **(F)** Variability of the amplitude of the eyelid startle responses as a function of prepulse intensity for children with autism and NT children. For all prepulse intensities the group of children with autism had a higher variability than NT children. **(G)** Box and density plots showing the percentage of PPI in trials with a high (25% of pulse) prepulse intensity. Subjects with a median startle amplitude for pulse trials < 0.075 were classified as non-responders and excluded from this plot. In more than 50 percent of the trials there was no decrease, but even an increase, in the startle response amplitude in children with autism. **(H)** Increased variability in the amplitude of the eyelid startle response during trials with a medium prepulse intensity in children with autism compared to NT children. Colored shading in panel A and error bars in panels C, F, G indicate 95% confidence intervals. Significance levels: * p < 0.05, ** p < 0.01, *** p < 0.001. All data and statistical analyses are available in **supplementary tables 1and 2**. **Supplementary** Figure 4 shows typical examples of raw eyelid movement profiles during PPI.

### Long-term habituation (long-term HAB)

Over the course of the 15-minute PPI test, children with autism showed significant habituation. There was a decrease in startle amplitude from 0.38 (±0.36) in the first two trials to 0.28 (±0.32) in trials 7-8 (**figure 2B, C**). In neurotypical children, while there was a decrease in startle amplitude from 0.30 (±0.34) in trials 1-2 compared to 0.26 (±0.29) in trials 7-8, this decrease was not significant (**supplementary table 2A**). While the main effect of trial number was significant (F_3,2233_=9.12, p<0.0001), the diagnosis (F_1,381_=3.27, p=0.071) and trial number * diagnosis interaction effects (F_3,2233_=1.95, p=0.12) were not significant. Sex was a significant confounder (F_1,380_=6.46, p=0.011), however, age was not (**supplementary table 2A)**. The significant decrease in startle amplitude over the course of the PPI test in autistic but not NT children is further supported by the linear regression analysis of ASR amplitudes (**supplementary fig. 3, supplementary table 2B**). The mean Pearson correlation coefficient for children with autism was -0.16 (±0.47), in comparison to -0.04 (±0.42) for NT children. There was a significant effect of diagnosis on the Pearson correlation coefficient (b = -0.12, t_381_=-2.53, CI [-0.21 - -0.03], p=0.048). This was only the case for pulse-only trials. No significant correlation was found for children with autism or neurotypical children for any of the prepulse categories (**supplementary fig. 3, supplementary table 2B)**.

### Prepulse inhibition (PPI)

Children with autism demonstrated reduced levels of PPI and increased variability in their startle eyelid responses **(figure 2D-H, supplementary fig. 4, supplementary table 1A, B)**. In neurotypical children, we found a statistically significant reduction in the amplitude of the eyelid startle response as the prepulse intensity increased, whereby the normalized eyelid closure (NEC) progressively decreased from 0.29 (±0.31) in trials without a prepulse to 0.25 (±0.30) in trials with a prepulse intensity at 10% of the pulse intensity (t_4164_=3.64, p=0.0006) and 0.22 (±0.28) in trials with the maximum prepulse intensity (t_4164_=7.49, p<0.0001). In contrast, children with autism did not exhibit a corresponding amplitude reduction for the prepulse intensity at 10% of the pulse (mean = 0.33 (±0.34) for pulse only trials versus 0.32 (±0.35) for prepulse 10% trials (t_5868_=1.74, p=0.16). However, they did show a significant decrease from the pulse trials to a mean of 0.30 (±0.34) for trials with the maximum prepulse intensity (t_5868_=4.02, p=0.0003).

We found a statistically significant effect of diagnosis for the trials with prepulse intensities at 10% (b = 0.07, t_380_=2.82, CI [0.02 - 0.11], p=0.010) and 25% (b = 0.09, t_376_=3.73, CI [0.04 - 0.13], p=0.001). There were no significant confounding effects of age or sex for any of the prepulse intensities. Interestingly, children with autism exhibited significantly increased variability in response amplitudes for all prepulse intensities and neither sex nor age were significant confounders (**figure 2F, H, supplementary table 1A**).

### Short-term habituation (short-term HAB)

In both the rhythmic and random stimulus patterns, we observed no decrease (i.e., habituation) in the mean amplitude of eyelid startle responses over the course of the six startle pulses. This lack of habituation was consistent across both children with autism and neurotypical children and neither sex nor age were significant confounders (**figure 3A, B**, **supplementary fig. 5, supplementary table 3A**). Notably, reflexive startle amplitudes tended to increase slightly after the first pulse in the group with autism, with a statistically significant startle amplitude increase at pulse 3 in the random stimulus pattern (pulse 1 vs. pulse 3: t_16767_=-3.19, p=0.0070).

**Figure 3.**
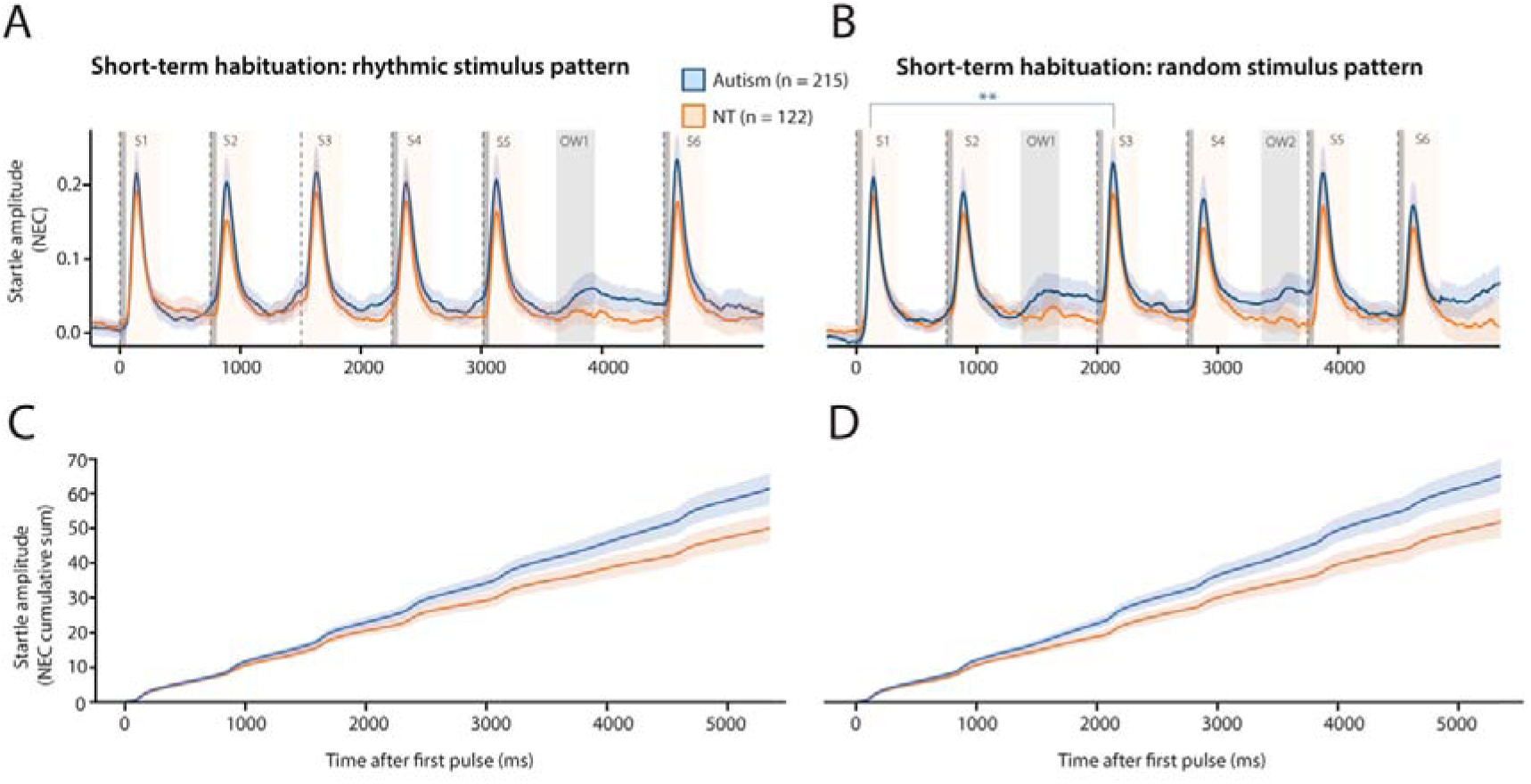
| Short-term startle habituation and anticipatory eyeblinks in a rhythmic and random stimulus pattern (A-B) Group averaged plots of eyelid movement over a duration of five seconds during which eyelid startle responses were evoked by the presentation of six white noise pulses in either a rhythmic (**A**) or a more random (**B**) stimulus pattern. Each 15- minute smartphone experiment consisted of ten rhythmic and ten random trials. Onset of the white noise pulses is indicated with the vertical dashed lines, duration of pulses (50 ms each) by the gray vertical columns. The magnitude of the startle pulse-evoked eyelid movements, expressed as normalized eyelid closure (NEC), was quantified in the ‘startle windows’ indicated by the yellow shaded areas S1 to S6. The magnitude of anticipatory eyeblink (AEB) responses around the moment of omitted stimuli was quantified in the gray-shaded observation windows (OW) 1 and 2. NT children showed no short-term habituation and children with autism even showed sensitization (i.e. startle responses becoming larger after the first pulse) in the random stimulus pattern. **(C-D)** Group averaged plots showing the cumulative sum of eyelid movements shown in panels A and B. The cumulative sum was higher and more variable in children with autism compared to NT children during the rhythmic and random trials. Colored shading indicates 95% confidence intervals. Significance levels: * p < 0.05, ** p < 0.01, *** p < 0.001. All data and statistical analysesare available in **Supplementary Tables 3**. **Supplementary** Figure 5 shows typical examples of raw eyelid movement profiles during short-term HAB.

### Anticipatory eye blinks (AEB)

Here we looked at whether eye blinks were present during windows where stimuli were omitted. We did not find statistically significant differences in eyelid closure amplitude in any of the predefined observation windows for either the rhythmic or random pulse trains (**supplementary table 3B**). While sex was not a significant confounder, age was for observation window 1 in the rhythmic protocol (b = 0.01, t_332_=2.32, CI [0.002 - 0.02], p = 0.021) and random protocol (b = 0.01, t_334_=2.19, CI [0.001 - 0.02], p = 0.029) as well as for observation window 2 in the random protocol (b = 0.01, t_334_=3.46, CI [0.01 - 0.02], p = 0.0006). When quantifying short-term HAB in conjunction with AEBs, using the cumulative summation of the mean eyelid amplitude in the two short-term HAB protocols, we did see differences between children with autism and neurotypical children. The cumulative sum for the rhythmic protocol was significantly different (b = 18.06, t_333_=2.94, CI [5.97 - 30.15], p = 0.0035, **figure 3C, supplementary table 3C**) between autistic children (cumulative NEC across 10 trials=78.36 ±108.98) and neurotypical children (cumulative NEC across 10 trials =61.00 ±75.42). For the random stimulus pattern, the mean cumulative sum of the eyelid amplitude was significantly higher (b = 19.16, t_335_=2.99, CI [6.54 - 31.78], p = 0.0030, **figure 3D, supplementary table 3C**) in children with autism, (cumulative NEC=81.90 ±114.52), compared to typically developing children (cumulative NEC=63.92 ±83.17). Age was a significant confounder (b = 3.73, t_334_=2.43, CI [0.71 - 6.75], p = 0.016), however, the main effects of autism stayed significant in the model adjusted for age (b = 15.48, t_334_=2.37, CI [2.62 - 28.35], p = 0.019, **supplementary table 3C**). We also quantified the variability in the maximum cumulative sum of the eyelid closure amplitude for the rhythmic and random stimulus patterns (**supplementary table 3C)**. Children with autism showed significantly more variability (b = 32.91, t_335_= 6.60, CI [23.10 - 42.73], p <0.0001) in the maximum cumulative sum of eyelid closure amplitude (mean SD = 87.14 ±48.96) compared to neurotypical children (mean SD = 54.22 ±33.52) in the rhythmic stimulus pattern. The same was true for the random stimulus pattern (b = 28.91, t_335_= 5.36, CI [18.30 - 39.52], p < 0.0001) where autistic children had a mean SD of 89.55 (± 52.52) and neurotypical children had a mean SD of 60.64 (±37.30). For the rhythmic pattern, age was a significant confounder (b = 2.38 t_335_= 2.00, CI [0.04 - 4.73], p = 0.047), however, the effect of diagnosis remained significant (b = 30.61, t_335_= 6.00, CI [20.58 - 40.64], p < 0.0001).

### Screen avoidance

Autistic children avoided watching the smartphone’s screen significantly more than neurotypical children (odds ratio (OR) = 6.09, CI [4.16 - 8.93], p < 0.0001; **figure 4A, supplementary table 4**). On average, children with autism displayed this behavior in 9.94% (±11.56) of trials, while neurotypical children faced away in 3.66% (±7.76) of trials. Age was a significant confounder (OR = 0.84, CI [0.77 - 0.92], p = 0.001), however, the main effects of autism remained significant in the model adjusted for age (OR = 7.26, CI [4.91 - 10.75], p < 0.0001).

**Figure 4.**
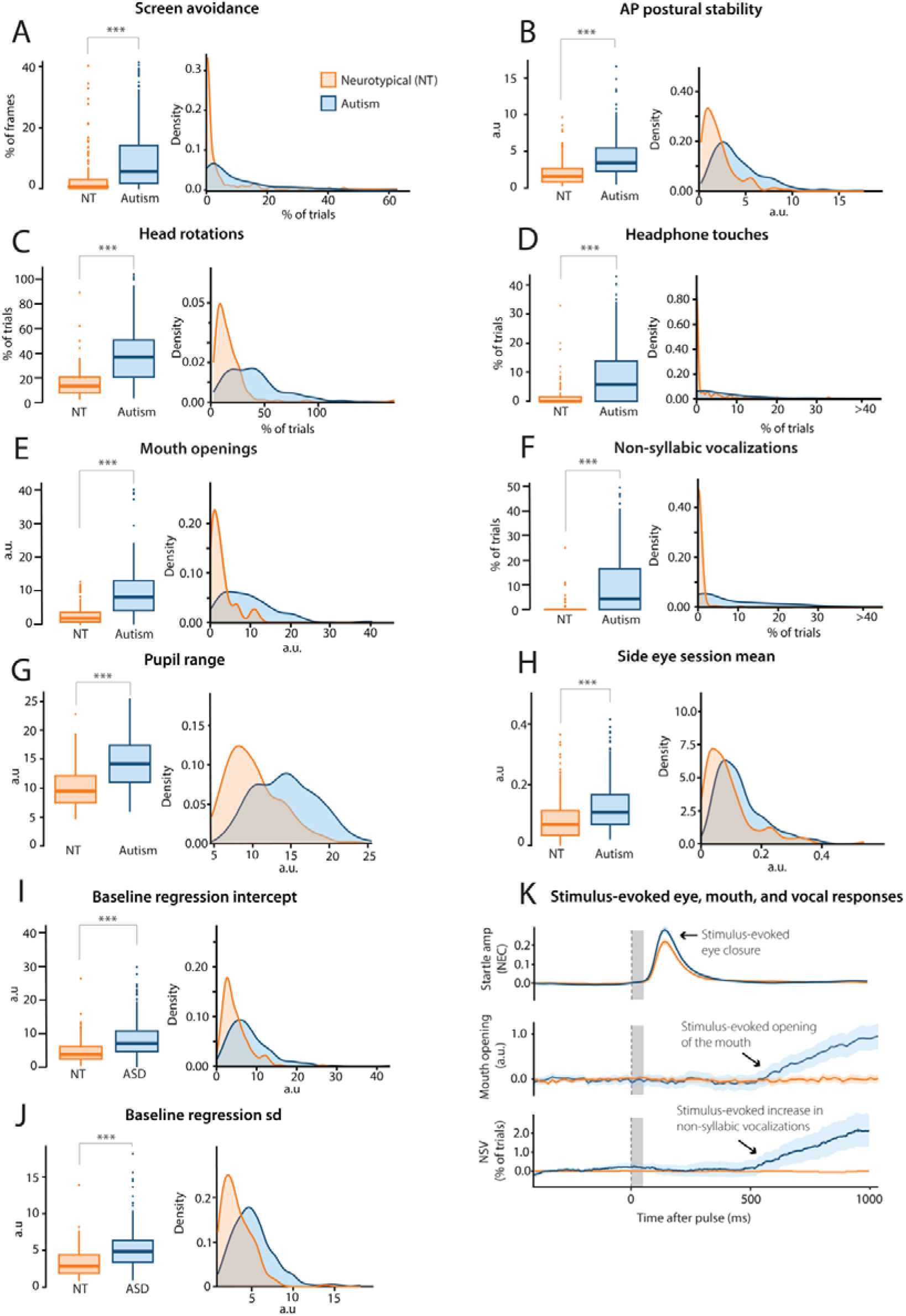
| Postural, head, facial, vocal and eyelid responses during the smartphone neurobehavioral evaluations differ between children with autism and neurotypical (NT) children. (A-J) Box plots and density plots illustrate increased response levels in children with autism (blue) compared to NT children (orange). A missing cap at the whiskers of the box plot means that the whiskers are not capturing the full data range, the range in density plots can therefore extend the range in the box plot. (K) Group averaged profiles of startle eyeblinks (normalized eyelid closure, NEC), and baseline-corrected mouth openings and non- syllabic vocalizations (NSV) around the onset of the auditory stimulus. Vertical dashed line indicates the onset of the auditory pulse. Gray vertical columns indicate the stimulus durations of 50 ms. The auditory stimulus evokes a startle eyeblink with a short latency to onset, followed by a mouth opening and vocalization with a longer latency to onset. Blue and orange shadings indicate the 95% confidence intervals. Significance levels: * p < 0.05, ** p < 0.01, *** p < 0.001. All data and statistical analyses are available in Supplementary Table 4. AP = Anterior posterior; SD = Standard deviation

### Anteroposterior postural stability

Children with autism exhibited significantly more anteroposterior postural movements during the experiments compared to their neurotypical counterparts (β = 0.48, CI [0.83 - 1.17], p<0.0001; **figure 4B, supplementary table 4**). Children with autism demonstrated a mean standard deviation of the facial bounding box size of 4.24 (±2.82), whereas neurotypical children exhibited a value of 2.11 (±1.77).

### Head rotations

Autistic children demonstrated significantly more head rotations compared to neurotypical children (β *=* 0.54, CI [0.95 - 1.28], p<0.0001; **figure 4C, supplementary 4**). Children with autism exhibited head rotations in 40.22% (±25.62) of trials while neurotypical children displayed head rotations in 16.62% (±16.76) of trials.

### Headphone touches

Children with autism displayed significantly more headphone touches during the experiments (OR = 16.67, CI [10.28 - 27.04], p<0.0001; **figure 4D, supplementary table 4**). Children with autism displayed headphone touches in 11.31% (±16.74) of trials, contrasting with neurotypical children who exhibited this behavior in 1.87% (±5.97) of trials.

### Mouth openings

Autistic children displayed both an increased frequency and larger mouth openings compared to neurotypical children (β *=* 0.55, CI [0.97 - 1.30], p<0.0001; **figure 4E, K, supplementary table 4**). The mean mouth surface area for autistic children was 9.55 (±6.95) versus a mean of 2.96 (*±*3.02) in neurotypical children. In addition, we were able to elicit mouth opening with auditory stimuli in children with autism, a response not observed in neurotypical controls **(figure 4K)**.

### Vocalizations

Children with autism demonstrated a significantly higher percentage of non- syllabic vocalizations during the experiments (OR = 95.33, CI [45.44 - 199.97], p<0.0001; **figure 4F, supplementary table 4**). Notably, these vocalizations occurred spontaneously throughout the 15-minute experiment in addition to being elicited by auditory stimuli. As with mouth openings, these stimulus-elicited responses were not observed in neurotypical controls **(figure 4K)**. Non-syllabic vocalizations were observed in 11.16% (±15.21) of trials in autistic children, in contrast to 0.43% (±2.40) of trials in neurotypical children (**figure 4F**).

### Pupil range

Autistic children displayed significantly more horizontal pupil movements compared to neurotypical children (β = 0.47, CI [3.45 - 4.97], p<0.0001; **figure 4G, supplementary table 4).** In autistic children the mean horizontal pupil movement was 14.33 (± 4.04) versus a mean of 10.12 (± 3.48) in neurotypical children.

**Side eye:** Children with autism also tended to more frequently look in the opposite direction from where their head was turned, referred to in this study as “side-eyeing” (β = 0.27, CI [0.36 - 0.74], p<0.0001; **figure 4H, supplementary table 4).** The mean amount of side-eye in children with autism was 0.13 (±0.08) compared to 0.09 (±0.08) in neurotypical children.

### Baseline regression measures

More variation in baseline eyelid opening was observed in autistic children. Children with autism showed more variation in baseline eyelid opening in the early stages of the experiment (quantified by the baseline regression intercept; β = 0.36, CI [0.57 - 0.94], p<0.0001; **figure 4I, supplementary table 4**). The mean baseline regression intercept was 8.31 (±5.55) in autistic children compared to 4.77 (±3.51) in neurotypical children. This variation in baseline eyelid opening was also more variable in children with autism compared to neurotypical children (β *=* 0.40, CI [0.65 - 1.02], p<0.0001; **figure 4J, supplementary table 4** indicated by a higher baseline regression SD in autistic (mean = 5.23 (±2.60) compared to neurotypical children (mean = 3.27 (±1.85).

### Exploratory analyses

To explore whether the outcomes described above were indeed autism-related and not due to other diagnoses that frequently co-occur with autism, we performed sub-analyses for the effect of co-occurring conditions on outcome measures that differed significantly between neurotypical and autistic children. For these analyses we focused on the two most common co-occurring conditions in this sample, which were intellectual disability (ID) and attention-deficit hyperactivity disorder (ADHD) (Khachadourian et al., 2023). Information on co-occurring conditions was available for 92% of the children with autism (**supplementary table 5**). While there was a significant effect of the co-occurring conditions group on age (F_3,249_= 2.89, p = 0.036), none of the three co-occurring condition groups differed significantly in age from the group without co-occurring conditions (ADHD vs autism only: t_249_= -1.30, p = 0.196, ID vs autism only: t_249_= 1.94, p = 0.053, ADHD & ID vs autism only: t_249_= -0.46, p = 0.650).

The only co-occurring condition group to show a significant difference in the amplitude of the ASR compared to the group without co-occurring conditions was the group with both ADHD and ID (b = -0.10, t_214_ = -2.15, CI [-0.19 - -0.01], p = 0.033, **supplementary table 6A**). There was no significant effect of co-occurring condition on long-term HAB (**supplementary table 7)**; however, the group with autism and ADHD did not show a significant decrease in the amplitude of the response to the pulse over the course of the experiment, whereas the other co-occurring condition groups did.

In the prepulse trials with the prepulse intensity at 25% of the pulse, children with autism and ID showed a significantly smaller amplitude of NEC, compared to children with autism and no co-occurring conditions (b = -0.12, t_209_= -3.04, CI [-0.19 - -0.04], p=0.008, **supplementary table 6A**). Of the groups of children with autism, the group with ADHD only (F_3,717_= 2.66, p = 0.047) and ID only (F_3,1623_= 8.73, p <0.0001) showed a significant effect of the prepulse on eyelid closure amplitude, whereas children with both ADHD and ID and those without co-occurring conditions did not show significant PPI (**supplementary table 6B**).

There was no effect of co-occurring conditions on short-term habituation for both the rhythmic and random stimulus patterns (**supplementary table 8A, B**). Similarly, there was no effect of the co-occurring conditions on the cumulative sum of the mean NEC amplitude or the variability of the maximum cumulative sum in either of the two short-term HAB protocols (**supplementary table 9).**

Mouth openings was the only sensorimotor behavior where a significant difference was observed between children with autism with and without co-occurring conditions. Specifically, autistic children with both ADHD and ID showed significantly more mouth openings (mean = 12.66 ±8.13) compared to children with autism and no co-occurring conditions (mean = 8.40 ±5.60, β = 0.59, CI [0.24 - 0.85], p = 0.0056, **supplementary table 10**).

## Discussion

In this study we assessed the ability of smartphone-based neurobehavioral quantification of sensorimotor functioning to distinguish between children with and without autism. We successfully evaluated 431 children aged 3 to 12 years with the BlinkLab smartphone platform. In this large sample, we confirmed that autistic children showed diminished PPI levels compared to neurotypical children. This finding should, however, be considered alongside the potential effect of co-occurring conditions on PPI as children with autism and ID showed more PPI compared to children with autism only. When sex, which was a significant confounder, was accounted for, the amplitude of the ASR was higher in autistic compared to neurotypical children. Furthermore, children with autism showed more variability in eyelid closure in both PPI and HAB experiments, which was not better explained by age, sex or the presence of co-occurring conditions. Interestingly, children with autism showed significantly more long-term habituation of the ASR over the course of a PPI experiment compared to neurotypical children.

In addition, several of the spontaneous and evoked behavioral parameters acquired with our digital platform showed significant differences amongst the diagnostic groups. Specifically, non-syllabic vocalizations were more common in autistic children, aligning with prior research (Tenenbaum et al., 2020). Likewise, postural movements and mouth openings were increased in children with autism, as well as head rotations, headphone touches, and screen avoidance. These results are compatible with other studies using video presentations (Dawson et al., 2018; Krishnappa Babu et al., 2023; Martin et al., 2018). It is unlikely that these behaviors were driven by conditions that frequently occur with autism, as all but the mouth openings showed no differences between autistic children with and without co-occurring conditions.

This more dynamic and unstable spontaneous and reflexive sensorimotor behavior in autism may arise from multiple neural mechanisms. Possible subcortical mechanisms include increased variability in coordination signals arising from the cerebellum (Peter et al., 2016), alterations in the output of the periaqueductal gray (Bandler, 1982; Jürgens, 1994), and/or abnormal brainstem processing (Schmitt et al., 2014).

The current study is one of the first to exploit smartphone technology and computer vision analysis to study simple forms of learning behavior in autism, including PPI and habituation of acoustic startle responses. Similar to studies done with smaller numbers of subjects in a laboratory setting (Cheng et al., 2018; Dwyer et al., 2023; Madsen et al., 2014; McAlonan et al., 2002; Perry et al., 2007), we found lower levels of PPI in autism in terms of eyelid closure amplitude. In addition, we found that autistic children had increased variability in their response amplitudes in both PPI and HAB sessions, consistent with the findings of Bhaskaran and colleagues (Bhaskaran et al., 2023) and Haigh (Haigh, 2018). Likewise, autistic children showed both a higher cumulative sum of NEC amplitude and variability in the maximum cumulative sum of NEC amplitude in HAB experiments, suggesting not merely a hyper-responsiveness to repeated auditory stimuli but also more inconsistencies in this hyper-responsiveness. Together our data raise the possibility that variability in sensory motor processing rather than a simple hypo- or hyperresponsiveness to sensory stimuli may better explain autism-related sensory differences.

By using both rhythmic and random stimulus trains, we determined the level of short-term habituation. Neither neurotypical nor children with autism showed decrements in their responses following repeated stimulation with the same stimulus. This coincides with a lab-based study (Muenssinger et al., 2013), which found weak or even no habituation in children exposed to blocks of eight tones separated by 300ms. Interestingly, children with autism did show significant habituation over a longer time period, i.e., over the course of the entire PPI experiment. The divergence in the NEC amplitude between neurotypical children and children with autism is significant for early trials, and then decreases over the course of subsequent trials, which indicates that the NEC amplitude in early trials possibly drives the observed difference in habituation. In contrast, in the short-term HAB paradigm, autistic children, unlike neurotypical children, showed a slight increase in their startle amplitudes after the first 2 pulses in the random stimulus train, corresponding with previous findings (Madsen et al., 2014) and potentially reflecting stronger sensitization.

In summary, we found that a smartphone platform can provide rapid evaluation of differences in sensorimotor function between neurotypical and autistic children. While our data-driven analyses reveal highly significant general sensorimotor behavioral differences between neurotypical children and those with autism, they lack the specificity of neurobehavioral reflex-based measures. Thus, combining these distinct potential biomarkers may hold the best potential in differentiating between individuals with and without autism.

### Study strengths and limitations

Strengths of our study lie in evaluating the BlinkLab app in an everyday environment with a sample of unmedicated children identified through a multi-center design. Existing technologies for autism often conduct research and validation studies in predominantly white, Western, male populations with direct access to optimal medical care (Ponzo et al., 2023). By conducting our study in a more sex-balanced, and geographically underrepresented sample, we aim to reduce existing barriers and enhance the technology’s applicability on a global scale (Dawson et al., 2023; Kirby et al., 2022).

Smartphone-mediated neurobehavioral assessments present novel technical challenges, particularly when assessing young children. These challenges include issues such as experimental disruptions caused by unreliable Wi-Fi networks, accidental or deliberate disconnection of headphones or adjustments to the volume settings, improper positioning of the smartphone, and instances where the child covers the camera. These issues are potentially addressable with extensions to the software platform’s user interface and feedback mechanisms.

This study was limited by the lack of detailed clinical characterization of the participants and the over-representation of children with autism compared to the global prevalence of the disorder. Future work should ensure neurotypical children are also evaluated by medical professions and could investigate potential correlations between symptom severity and smartphone neurobehavioral measures. Stratifying children by level of functioning was, however, beyond the scope of this study where the primary aim was to first assess whether smartphone-based neurobehavioral measures could distinguish differences in sensorimotor functioning.

Expanding this retrospective case-control study to a pre-registered prospective clinical trial would be a next step for future work to evaluate the potential diagnostic accuracy of a smartphone tool for autism. Finally, repeated measurements from a cohort will allow the effects of behavioral and pharmacological interventions on sensorimotor function to be quantified longitudinally.

## Conclusion

Our smartphone-based testing addresses a gap in existing mobile platforms by focusing on sensorimotor function through neurobehavioral evaluations, which until now have been demonstrated primarily in academic settings in autistic individuals and animal models. Smartphone-based tests do not require verbal or social interaction, closing research gaps arising from the historical exclusion of non-verbal individuals with autism from research, and complementing other testing platforms that measure social interactions using human scoring of behavior. Our results serve as a starting point from which to investigate the diagnostic accuracy of a smartphone tool in a future prospective clinical trial.

## Data Availability

The datasets generated or analyzed during the study are available from the corresponding author on reasonable request.

## Acknowledgements

We extend our appreciation to the families involved in our study for their commitment and contribution to advancing knowledge of ASD. We wish to express our gratitude to Dr. Mohammed El Azami, coordinator of Mohammed V Foundation for Solidarity for assisting us implement this study at the various centers and to the dedicated team in Morocco and their diligence in testing and supporting the participants, the contributors are Hicham El Idrissi, Mohammed Hamel, Tayouby Aziz, Lahniche Aziz, Rhazil Hasnaa, Mourad Gadi, Driouach Hassnae, Belhaj Naila, Boutahiri Abdelhak, El Mansour Khadija, Msaidi Abdellah, Mohammed Zouggari, Noureddin Oukoodda, Rabi Nouh, Mohammed Naim, Fouad Ichir, Fadwa Gharrabou, Dalal Rhamouch, Jahjah Hadda, Boulgana Nouama, Bouzzit Amina, Fadoua Ouaziz, Aicha Aziz, Rachida Badrkane, Rania Triki, Nabil Belhoucine, Azzi Lakbira, Rajaa Baymik, and Alaoui Ghita. We thank Danique Paula for technical assistance. We thank AWS Activate and NVIDIA inception for providing complimentary cloud credits. CIDZ is funded by Medical Neuro-Delta (MD 01092019-31082023), INTENSE LSH-NWO (TTW/00798883), ERC-adv (GA-294775) and ERC-POC (nrs. 737619 and 768914), as well as the Dutch NWO Gravitation Program, Dutch Brain Interface Initiative (DBI2 grant no. 024.005.022).

## Contributors

HJB, SSHW, AU, CPB, SKEK conceived and designed the study. HJB, SS, AEI, SSHW, AU, CPB, SKEK, KG, LEMR, SL were responsible for the methodology and HJB, SSHW, AEI, LEMR, SKEK, KG, SM, TFS, VCA carried out the investigation. The analysis was conducted by HJB, CPB, SKEK, SS, KG, SL, MO and visualization was done by HJB, SKEK and KG. HJB and SSHW acquired funding for this project. Project administration was carried out by HJB, SS, AEI, VCA, LEMR, KG, KB, SAB. HJB, AU, SKEK, CIDZ, SSHW, MO were responsible for supervision. The original draft of the manuscript was written by KG, HJB, MO and reviewed and edited by HJB, KG, MO, SL, CJ, SS, CPB, CIDZ, SSHW, AEI, LEMR, ES. All authors had the final responsibility for the decision to submit for publication.

Initials: H.J. Boele (HJB), S.S.-H. Wang (SSHW), A. Uvarov (AU), C.P. Boele (CPB), S.K.E. Koekkoek (SKEK), S. Sherry (SS), A. El Idrissi (AEI), K. Gultig (KG), L.E.M. Roggeveen (LEMR), Sander Lindeman (SL), S. Milosevska (SM), C. Jung (CJ), E. Sefik (ES), Ting Fang Soong (TFS), K. Benhassan (KB), S. Ait BenAli (SAB), V. Carpio-Aras (VCA), C.I. De Zeeuw, (CIDZ), Myrthe Ottenhoff (MO).

## Ethics

The study was performed in accordance with relevant guidelines and regulations and was reviewed and approved by the institutional review boards of Princeton University (#13943) and the Faculté de Médecine et de Pharmacie de Marrakech in Morocco (#23/2022).

## Patient consent

The researcher and parents of the children pictured provided informed consent for publication of these images in an online open-access publication.

## References

Abel, K., Waikar, M., Pedro, B., Hemsley, D., & Geyer, M. (1998). Repeated testing of prepulse inhibition and habituation of the startle reflex: A study in healthy human controls. Journal of Psychopharmacology, 12(4), 330–337. 10.1177/026988119801200402

American Psychiatric Association, D. S. M. T. F., & American Psychiatric Association. (2013). Diagnostic and statistical manual of mental disorders: DSM-5 (5th ed., Vol. 5). American psychiatric association. 10.1002/aur.3022

Bandler, R. (1982). Induction of ‘page’ following microinjections of glutamate into midbrain but not hypothalamus of cats. Neuroscience Letters, 30(2), 183–188. 10.1016/0304-3940(82)90294-4

Bhaskaran, A. A., Gauvrit, T., Vyas, Y., Bony, G., Ginger, M., & Frick, A. (2023). Endogenous noise of neocortical neurons correlates with atypical sensory response variability in the Fmr1−/y mouse model of autism. Nature Communications, 14(1), Article 1. 10.1038/s41467-023-43777-z

Boele, H. J., Jung, C., Sherry, S., Roggeveen, L. E. M., Dijkhuizen, S., Öhman, J., Abraham, E., Uvarov, A., Boele, C. P., Gultig, K., Rasmussen, A., Vinueza-Veloz, M. F., Medina, J. F., Koekkoek, S. K. E., De Zeeuw, C. I., & Wang, S. S.-H. (2023). Accessible and reliable neurometric testing in humans using a smartphone platform. Scientific Reports, 13(1), Article 1. 10.1038/s41598-023-49568-2

Braff, D. L., Geyer, M. A., & Swerdlow, N. R. (2001). Human studies of prepulse inhibition of startle: Normal subjects, patient groups, and pharmacological studies. Psychopharmacology, 156(2/3), 234. 10.1007/s002130100810

Campbell, K., Carpenter, K. L., Hashemi, J., Espinosa, S., Marsan, S., Borg, J. S., Chang, Z., Qiu, Q., Vermeer, S., Adler, E., Tepper, M., Egger, H. L., Baker, J. P., Sapiro, G., & Dawson, G. (2019). Computer vision analysis captures atypical attention in toddlers with autism. Autism: The International Journal of Research and Practice, 23(3), 619– 628. 10.1177/1362361318766247

Cheng, C.-H., Chan, P.-Y. S., Hsu, S.-C., & Liu, C.-Y. (2018). Meta-analysis of sensorimotor gating in patients with autism spectrum disorders. Psychiatry Research, 262, 413–419. 10.1016/j.psychres.2017.09.016

Coll, S.-M., Foster, N. E. V., Meilleur, A., Brambati, S. M., & Hyde, K. L. (2020). Sensorimotor skills in autism spectrum disorder: A meta-analysis. Research in Autism Spectrum Disorders, 76, 101570. 10.1016/j.rasd.2020.101570

Dawson, G., Campbell, K., Hashemi, J., Lippmann, S. J., Smith, V., Carpenter, K., Egger, H., Espinosa, S., Vermeer, S., Baker, J., & Sapiro, G. (2018). Atypical postural control can be detected via computer vision analysis in toddlers with autism spectrum disorder. Scientific Reports, 8, 17008. 10.1038/s41598-018-35215-8

Dawson, G., Rieder, A. D., & Johnson, M. H. (2023). Prediction of autism in infants: Progress and challenges. The Lancet Neurology, 22(3), 244–254. 10.1016/S1474-4422(22)00407-0

Denisova, K., & Wolpert, D. M. (2024). Sensorimotor variability distinguishes early features of cognition in toddlers with autism. iScience, 27(9), 110685. 10.1016/j.isci.2024.110685

Doornaert, E. E., Mohamad, A. E.-C., Johal, G., Allman, B. L., Möhrle, D., & Schmid, S. (2024). Not a Deficit, Just Different: Prepulse Inhibition Disruptions in Autism Depend on Startle Stimulus Intensities. eNeuro, 11(9), ENEURO.0179-24.2024. 10.1523/ENEURO.0179-24.2024

Durkin, M. S., Elsabbagh, M., Barbaro, J., Gladstone, M., Happe, F., Hoekstra, R. A., Lee, L.-C., Rattazzi, A., Stapel-Wax, J., Stone, W. L., Tager-Flusberg, H., Thurm, A., Tomlinson, M., & Shih, A. (2015). Autism screening and diagnosis in low resource settings: Challenges and opportunities to enhance research and services worldwide. Autism Research, 8(5), 473–476. 10.1002/aur.1575

Dwyer, P., Williams, Z. J., Vukusic, S., Saron, C. D., & Rivera, S. M. (2023). Habituation of auditory responses in young autistic and neurotypical children. Autism Research, 16(10), 1–21. 10.1002/aur.3022

El-Cheikh Mohamad, A., Möhrle, D., Haddad, F. L., Rose, A., Allman, B. L., & Schmid, S. (2023). Assessing the Cntnap2 knockout rat prepulse inhibition deficit through prepulse scaling of the baseline startle response curve. Translational Psychiatry, 13(1), 1–11. 10.1038/s41398-023-02629-6

Franz, L., Goodwin, C. D., Rieder, A., Matheis, M., & Damiano, D. L. (2022). Early intervention for very young children with or at high likelihood for autism spectrum disorder: An overview of reviews. Developmental Medicine and Child Neurology, 64(9), 1063–1076. 10.1111/dmcn.15258

Geyer, M. A., Krebs-Thomson, K., Braff, D. L., & Swerdlow, N. R. (2001). Pharmacological studies of prepulse inhibition models of sensorimotor gating deficits in schizophrenia: A decade in review. Psychopharmacology, 156(2–3), 117–154. 10.1007/s002130100811

Graham, F. K. (1975). The More or Less Startling Effects of Weak Prestimulation. Psychophysiology, 12(3), 238–248. 10.1111/j.1469-8986.1975.tb01284.x

Haigh, S. M. (2018). Variable sensory perception in autism. European Journal of Neuroscience, 47(6), 602–609. 10.1111/ejn.13601

Hannant, P., Cassidy, S., Tavassoli, T., & Mann, F. (2016). Sensorimotor Difficulties Are Associated with the Severity of Autism Spectrum Conditions. Frontiers in Integrative Neuroscience, 10. 10.3389/fnint.2016.00028

Jürgens, U. (1994). The role of the periaqueductal grey in vocal behaviour. Behavioural Brain Research, 62(2), 107–117. 10.1016/0166-4328(94)90017-5

Kanne, S. M., & Bishop, S. L. (2021). Editorial Perspective: The autism waitlist crisis and remembering what families need. Journal of Child Psychology and Psychiatry, and Allied Disciplines, 62(2), 140–142. 10.1111/jcpp.13254

Khachadourian, V., Mahjani, B., Sandin, S., Kolevzon, A., Buxbaum, J. D., Reichenberg, A., & Janecka, M. (2023). Comorbidities in autism spectrum disorder and their etiologies. Translational Psychiatry, 13(1), 1–7. 10.1038/s41398-023-02374-w

Kirby, A. V., Bilder, D. A., Wiggins, L. D., Hughes, M. M., Davis, J., Hall-Lande, J. A., Lee, L.-C., McMahon, W. M., & Bakian, A. V. (2022). Sensory features in autism: Findings from a large population-based surveillance system. Autism Research: Official Journal of the International Society for Autism Research, 15(4), 751–760. 10.1002/aur.2670

Koch, M. (1999). The neurobiology of startle. Progress in Neurobiology, 59(2), 107–128. 10.1016/S0301-0082(98)00098-7

Kohl, S., Wolters, C., Gruendler, T. O. J., Vogeley, K., Klosterkötter, J., & Kuhn, J. (2014). Prepulse Inhibition of the Acoustic Startle Reflex in High Functioning Autism. PLOS ONE, 9(3), e92372. 10.1371/journal.pone.0092372

Krishnappa Babu, P. R., Di Martino, J. M., Chang, Z., Perochon, S., Aiello, R., Carpenter, K. L. H., Compton, S., Davis, N., Franz, L., Espinosa, S., Flowers, J., Dawson, G., & Sapiro, G. (2023). Complexity analysis of head movements in autistic toddlers. Journal of Child Psychology and Psychiatry, 64(1), 156–166. 10.1111/jcpp.13681

Kumari, V., Zachariah, E., Galea, A., Mehrotra, R., Taylor, D., & Sharma, T. (2001). Effects of procyclidine on prepulse inhibition of the acoustic startle response in healthy human volunteers. Psychopharmacology, 154(3), 221–229. 10.1007/s002130000656

López-Schier, H. (2019). Neuroplasticity in the acoustic startle reflex in larval zebrafish. Current Opinion in Neurobiology, *54*, 134–139. 10.1016/j.conb.2018.10.004

Madsen, G. F., Bilenberg, N., Cantio, C., & Oranje, B. (2014). Increased prepulse inhibition and sensitization of the startle reflex in autistic children. Autism Research: Official Journal of the International Society for Autism Research, 7(1), 94–103. 10.1002/aur.1337

Martin, K. B., Hammal, Z., Ren, G., Cohn, J. F., Cassell, J., Ogihara, M., Britton, J. C., Gutierrez, A., & Messinger, D. S. (2018). Objective measurement of head movement differences in children with and without autism spectrum disorder. Molecular Autism, 9(1), 14. 10.1186/s13229-018-0198-4

McAlonan, G. M., Daly, E., Kumari, V., Critchley, H. D., van Amelsvoort, T., Suckling, J., Simmons, A., Sigmundsson, T., Greenwood, K., Russell, A., Schmitz, N., Happe, F., Howlin, P., & Murphy, D. G. M. (2002). Brain anatomy and sensorimotor gating in Asperger’s syndrome. Brain, 125(7), 1594–1606. 10.1093/brain/awf150

Muenssinger, J., Stingl, K. T., Matuz, T., Binder, G., Ehehalt, S., & Preissl, H. (2013). Auditory habituation to simple tones: Reduced evidence for habituation in children compared to adults. Frontiers in Human Neuroscience, 7. 10.3389/fnhum.2013.00377

Oranje, B., Lahuis, B., van Engeland, H., Jan van der Gaag, R., & Kemner, C. (2013). Sensory and sensorimotor gating in children with multiple complex developmental disorders (MCDD) and autism. Psychiatry Research, 206(2), 287–292. 10.1016/j.psychres.2012.10.014

Ornitz, E. M., Lane, S. J., Sugiyama, T., & de Traversay, J. (1993). Startle modulation studies in autism. Journal of Autism and Developmental Disorders, 23(4), 619–637. 10.1007/BF01046105

Patterson, J. W., Armstrong, V., Duku, E., Richard, A., Franchini, M., Brian, J., Zwaigenbaum, L., Bryson, S. E., Sacrey, L.-A. R., Roncadin, C., & Smith, I. M. (2022). Early trajectories of motor skills in infant siblings of children with autism spectrum disorder. Autism Research, 15(3), 481–492. 10.1002/aur.2641

Perry, W., Minassian, A., Lopez, B., Maron, L., & Lincoln, A. (2007). Sensorimotor gating deficits in adults with autism. Biological Psychiatry, 61(4), 482–486. 10.1016/j.biopsych.2005.09.025

Peter, S., ten Brinke, M. M., Stedehouder, J., Reinelt, C. M., Wu, B., Zhou, H., Zhou, K., Boele, H.-J., Kushner, S. A., Lee, M. G., Schmeisser, M. J., Boeckers, T. M., Schonewille, M., Hoebeek, F. E., & De Zeeuw, C. I. (2016). Dysfunctional cerebellar Purkinje cells contribute to autism-like behaviour in Shank2-deficient mice. Nature Communications, 7(1), Article 1. 10.1038/ncomms12627

Pilz, P. K. D., & Schnitzler, H.-U. (1996). Habituation and Sensitization of the Acoustic Startle Response in Rats: Amplitude, Threshold, and Latency Measures. Neurobiology of Learning and Memory, 66(1), 67–79. 10.1006/nlme.1996.0044

Ponzo, S., May, M., Tamayo-Elizalde, M., Bailey, K., Shand, A. J., Bamford, R., Multmeier, J., Griessel, I., Szulyovszky, B., Blakey, W., Valentine, S., & Plans, D. (2023). App Characteristics and Accuracy Metrics of Available Digital Biomarkers for Autism: Scoping Review. JMIR mHealth and uHealth, 11, e52377. 10.2196/52377

Schmitt, L. M., Cook, E. H., Sweeney, J. A., & Mosconi, M. W. (2014). Saccadic eye movement abnormalities in autism spectrum disorder indicate dysfunctions in cerebellum and brainstem. Molecular Autism, 5(1), 47. 10.1186/2040-2392-5-47

Simons-Weidenmaier, N. S., Weber, M., Plappert, C. F., Pilz, P. K., & Schmid, S. (2006). Synaptic depression and short-term habituation are located in the sensory part of the mammalian startle pathway. BMC Neuroscience, 7(1), 38. 10.1186/1471-2202-7-38

Swerdlow, N. R., Braff, D. L., & Geyer, M. A. (1999). Cross-species Studies of Sensorimotor Gating of the Startle Reflex. Annals of the New York Academy of Sciences, *877*(1 ADVANCING FRO), 202–216. 10.1111/j.1749-6632.1999.tb09269.x

Takahashi, H., Komatsu, S., Nakahachi, T., Ogino, K., & Kamio, Y. (2016). Relationship of the Acoustic Startle Response and Its Modulation to Emotional and Behavioral Problems in Typical Development Children and Those with Autism Spectrum Disorders. Journal of Autism and Developmental Disorders, 46(2), 534–543. 10.1007/s10803-015-2593-4

Takahashi, H., Nakahachi, T., Stickley, A., Ishitobi, M., & Kamio, Y. (2017). Stability of the acoustic startle response and its modulation in children with typical development and those with autism spectrum disorders: A one-year follow-up. Autism Research, 10(4), 673–679. 10.1002/aur.1710

Tenenbaum, E. J., Carpenter, K. L. H., SabatoslJDeVito, M., Hashemi, J., Vermeer, S., Sapiro, G., & Dawson, G. (2020). A SixlJMinute Measure of Vocalizations in Toddlers with Autism Spectrum Disorder. Autism Research, 13(8), 1373–1382. 10.1002/aur.2293

Travers, B. G., Bigler, E. D., Duffield, T. C., Prigge, M. D. B., Froehlich, A. L., Lange, N., Alexander, A. L., & Lainhart, J. E. (2017). Longitudinal development of manual motor ability in autism spectrum disorder from childhood to mid-adulthood relates to adaptive daily living skills. Developmental Science, 20(4). 10.1111/desc.12401

West, K. L. (2019). Infant Motor Development in Autism Spectrum Disorder: A Synthesis and Meta-analysis. Child Development, 90(6), 2053–2070. 10.1111/cdev.13086

Wilson, R. B., Vangala, S., Elashoff, D., Safari, T., & Smith, B. A. (2021). Using Wearable Sensor Technology to Measure Motion Complexity in Infants at High Familial Risk for Autism Spectrum Disorder. *Sensors (Basel*, Switzerland), 21(2), 616. 10.3390/s21020616

Yuhas, J., Cordeiro, L., Tassone, F., Ballinger, E., Schneider, A., Long, J. M., Ornitz, E. M., & Hessl, D. (2011). Brief Report: Sensorimotor Gating in Idiopathic Autism and Autism Associated with Fragile X Syndrome. Journal of Autism and Developmental Disorders, 41(2), 248–253. 10.1007/s10803-010-1040-9

Zhao, Z., Zhu, Z., Zhang, X., Tang, H., Xing, J., Hu, X., Lu, J., Peng, Q., & Qu, X. (2021). Atypical Head Movement during Face-to-Face Interaction in Children with Autism Spectrum Disorder. Autism Research: Official Journal of the International Society for Autism Research, 14(6), 1197–1208. 10.1002/aur.2478

Zhao, Z., Zhu, Z., Zhang, X., Tang, H., Xing, J., Hu, X., Lu, J., & Qu, X. (2022). Identifying Autism with Head Movement Features by Implementing Machine Learning Algorithms. Journal of Autism and Developmental Disorders, 52(7), 3038–3049. 10.1007/s10803-021-05179-2

Zwaigenbaum, L., Bauman, M. L., Stone, W. L., Yirmiya, N., Estes, A., Hansen, R. L., McPartland, J. C., Natowicz, M. R., Choueiri, R., Fein, D., Kasari, C., Pierce, K., Buie, T., Carter, A., Davis, P. A., Granpeesheh, D., Mailloux, Z., Newschaffer, C., Robins, D., … Wetherby, A. (2015). Early Identification of Autism Spectrum Disorder: Recommendations for Practice and Research. Pediatrics, 136(Suppl 1), S10–S40. 10.1542/peds.2014-3667C

