## supplementary tables for "Neurobehavioral Assessment of Sensorimotor Function in Autism Using Smartphone Technology"

| **Stimulus type** | **NT (n = 147)** | **Autism (n = 236)** | **Model** | **Main effect** | **B** | **SE** | **CI low** | **CI high** | **T-statistic** | **Adjusted R^2^** | **P-value** | **Adjusted p-value** |
| --- | --- | --- | --- | --- | --- | --- | --- | --- | --- | --- | --- | --- |
|  | **Eyelid startle amplitude (mean NEC ± SD)** | |  |  |  |  |  |  |  |  |  |  |
| **Pulse only** | 0.29 (± 0.31) | 0.33 (± 0.34) | Diagnosis | Diagnosis | 0.05 | 0.02 | 0.00 | 0.09 | 1.82 | 0.004 | 0.070 | . |
|  |  |  | Diagnosis + sex | Diagnosis | 0.05 | 0.03 | 0.01 | 0.10 | 2.19 | 0.010 | 0.029 | . |
|  |  |  |  | Sex | -0.06 | 0.03 | -0.11 | -0.01 | -2.52 |  | 0.012 | . |
|  |  |  | Diagnosis + age | Diagnosis | 0.04 | 0.03 | -0.01 | 0.10 | 1.71 | 0.005 | 0.087 | . |
|  |  |  |  | Age | 0.00 | 0.01 | -0.01 | 0.01 | 0.10 |  | 0.92 | . |
| **PP 5% + Pulse** | 0.28 (± 0.30) | 0.32 (± 0.35) | Diagnosis | Diagnosis | 0.05 | 0.02 | 0.00 | 0.09 | 1.88 | 0.005 | 0.061 | 0.060 |
|  |  |  | Diagnosis + sex | Diagnosis | 0.05 | 0.02 | 0.00 | 0.10 | 2.03 | 0.006 | 0.043 | 0.043 |
|  |  |  |  | Sex | -0.03 | 0.03 | -0.08 | 0.02 | -1.11 |  | 0.27 | 0.80 |
|  |  |  | Diagnosis + age | Diagnosis | 0.06 | 0.03 | 0.01 | 0.11 | 2.18 | 0.007 | 0.030 | 0.030 |
|  |  |  |  | Age | -0.01 | 0.01 | -0.02 | 0.00 | -1.31 |  | 0.19 | 0.57 |
| **PP 10% + Pulse** | 0.25 (± 0.30) | 0.32 (± 0.35) | Diagnosis | Diagnosis | 0.07 | 0.02 | 0.02 | 0.11 | 2.82 | 0.010 | 0.005 | 0.010 |
|  |  |  | Diagnosis + sex | Diagnosis | 0.07 | 0.02 | 0.02 | 0.12 | 2.87 | 0.010 | 0.004 | 0.009 |
|  |  |  |  | Sex | -0.01 | 0.02 | -0.06 | 0.04 | -0.51 |  | 0.61 | 0.80 |
|  |  |  | Diagnosis + age | Diagnosis | 0.08 | 0.02 | 0.03 | 0.13 | 3.09 | 0.012 | 0.002 | 0.004 |
|  |  |  |  | Age | -0.01 | 0.01 | -0.02 | 0.00 | -1.32 |  | 0.19 | 0.57 |
| **PP 25% + Pulse** | 0.22 (± 0.28) | 0.30 (± 0.34) | Diagnosis | Diagnosis | 0.09 | 0.02 | 0.04 | 0.13 | 3.73 | 0.017 | 0.000 | 0.001 |
|  |  |  | Diagnosis + sex | Diagnosis | 0.09 | 0.02 | 0.04 | 0.13 | 3.82 | 0.018 | 0.000 | 0.0005 |
|  |  |  |  | Sex | -0.02 | 0.02 | -0.07 | 0.03 | -0.86 |  | 0.39 | 0.80 |
|  |  |  | Diagnosis + age | Diagnosis | 0.09 | 0.02 | 0.05 | 0.14 | 3.93 | 0.019 | 0.000 | 0.0003 |
|  |  |  |  | Age | -0.01 | 0.01 | -0.02 | 0.00 | -1.24 |  | 0.22 | 0.57 |
|  | **Variability in eyelid startle amplitude (mean NEC ± SD)** | |  |  |  |  |  |  |  |  |  |  |
| **Pulse only** | 0.20 (± 0.11) | 0.25 (± 0.12) | Diagnosis | Diagnosis | 0.04 | 0.01 | 0.02 | 0.07 | 3.76 | 0.033 | 0.0002 | 0.001 |
|  |  |  | Diagnosis + sex | Diagnosis | 0.04 | 0.01 | 0.02 | 0.07 | 3.66 | 0.031 | 0.0003 | 0.001 |
|  |  |  |  | Sex | 0.00 | 0.01 | -0.02 | 0.03 | 0.38 | 0.031 | 0.71 | 1.000 |
|  |  |  | Diagnosis + age | Diagnosis | 0.04 | 0.01 | 0.02 | 0.07 | 3.57 | 0.031 | 0.0004 | 0.001 |
|  |  |  |  | Age | 0.00 | 0.00 | -0.01 | 0.01 | 0.14 | 0.031 | 0.89 | 1.000 |
| **PP 5% + Pulse** | 0.21 (± 0.10) | 0.25 (± 0.12) | Diagnosis | Diagnosis | 0.04 | 0.01 | 0.02 | 0.06 | 3.42 | 0.028 | 0.001 | 0.001 |
|  |  |  | Diagnosis + sex | Diagnosis | 0.04 | 0.01 | 0.02 | 0.06 | 3.27 | 0.027 | 0.001 | 0.001 |
|  |  |  |  | Sex | 0.01 | 0.01 | -0.01 | 0.03 | 0.73 | 0.027 | 0.47 | 1.000 |
|  |  |  | Diagnosis + age | Diagnosis | 0.04 | 0.01 | 0.02 | 0.07 | 3.60 | 0.029 | 0.0004 | 0.001 |
|  |  |  |  | Age | 0.00 | 0.00 | -0.01 | 0.00 | -1.14 | 0.029 | 0.26 | 1.000 |
| **PP 10% + Pulse** | 0.21 (± 0.11) | 0.26 (± 0.12) | Diagnosis | Diagnosis | 0.06 | 0.01 | 0.03 | 0.08 | 4.83 | 0.055 | <0.0001 | <0.0001 |
|  |  |  | Diagnosis + sex | Diagnosis | 0.06 | 0.01 | 0.03 | 0.08 | 4.72 | 0.053 | <0.0001 | <0.0001 |
|  |  |  |  | Sex | 0.00 | 0.01 | -0.02 | 0.03 | 0.27 | 0.053 | 0.79 | 1.000 |
|  |  |  | Diagnosis + age | Diagnosis | 0.06 | 0.01 | 0.03 | 0.08 | 4.73 | 0.053 | <0.0001 | <0.0001 |
|  |  |  |  | Age | 0.00 | 0.00 | -0.01 | 0.00 | -0.36 | 0.053 | 0.72 | 1.000 |
| **PP 25% + Pulse** | 0.20 (± 0.11) | 0.24 (± 0.11) | Diagnosis | Diagnosis | 0.04 | 0.01 | 0.02 | 0.07 | 3.66 | 0.032 | 0.0003 | 0.001 |
|  |  |  | Diagnosis + sex | Diagnosis | 0.04 | 0.01 | 0.02 | 0.06 | 3.46 | 0.032 | 0.001 | 0.001 |
|  |  |  |  | Sex | 0.01 | 0.01 | -0.01 | 0.04 | 1.04 | 0.032 | 0.30 | 1.000 |
|  |  |  | Diagnosis + age | Diagnosis | 0.04 | 0.01 | 0.02 | 0.07 | 3.33 | 0.030 | 0.001 | 0.001 |
|  |  |  |  | Age | 0.00 | 0.00 | 0.00 | 0.01 | 0.60 | 0.030 | 0.55 | 1.000 |

**Table S1A | Prepulse inhibition of the acoustically evoked eyelid startle response in children with autism compared to neurotypical children.**

All statistical comparisons done using Linear Mixed-Effect Models.

P-value adjustments: Bonferroni-Holm method for three tests for eyelid startle amplitude comparisons and for four tests for variability in eyelid startle amplitude comparisons.

NT = Neurotypical, NEC = Normalized Eyelid Closure, SD = Standard Deviation, SE = Standard Error, CI = 95% Confidence Interval, PP = prepulse.

| **ANOVA on LME for eyelid startle amplitude** | | | | |
| --- | --- | --- | --- | --- |
|  | | | **Main effect trial type** | |
|  |  |  | **F-test** | **P-value** |
| **NT (n = 147)** | | | F_3,4164_ = 22.14 | <0.0001 |
| **Autism (n = 236)** | | | F_3,5868_ = 5.47 | 0.0009 |
| **Pairwise differences** | | | | |
|  | **Estimate** | **T-ratio** | **P-value** | **Adjusted**  **p-value** |
| **NT** |  | | | |
| **(Pulse) vs. (PP 5% + Pulse)** | 0.02 | 1.10 | 0.27 | 0.27 |
| **(Pulse) vs. (PP 10% + Pulse)** | 0.04 | 3.64 | 0.0003 | 0.0006 |
| **(Pulse) vs. (PP 25% + Pulse)** | 0.07 | 7.49 | <0.0001 | <0.0001 |
| **Autism** |  | | | |
| **(Pulse) vs. (PP 5% + Pulse)** | 0.02 | 1.56 | 0.12 | 0.16 |
| **(Pulse) vs. (PP 10% + Pulse)** | 0.02 | 1.74 | 0.082 | 0.16 |
| **(Pulse) vs. (PP 25% + Pulse)** | 0.04 | 4.02 | 0.0001 | 0.0003 |

**Table S1B | Within group effect of trial type for prepulse inhibition of the acoustically evoked eyelid startle response in autistic and neurotypical children.**

All statistical comparisons done using ANOVA on Linear Mixed-Effect Model.

P-value adjustments: Bonferroni-Holm method for three tests.

LME = Linear Mixed-Effect Model, NT = Neurotypical, PP = prepulse.

|  | **Eyelid startle amplitude (mean NEC ± SD)** | | | | | | | | **Model** | **Main effect** | **F-test** | **P-value** |
| --- | --- | --- | --- | --- | --- | --- | --- | --- | --- | --- | --- | --- |
| **Trial number** | **NT (n = 147)** | | | | **Autism ( n = 236)** | | | | **Diagnosis x trial number** | Trial number | F_3.2233_ = 9.12 | < 0.0001 |
| **1-2** | 0.30 (± 0.34) | | | | 0.38 (± 0.36) | | | |  | Diagnosis | F_1.381_ = 3.27 | 0.071 |
| **3-4** | 0.30 (± 0.31) | | | | 0.35 (± 0.35) | | | |  | Diagnosis x trial number | F_3.2233_ = 1.95 | 0.12 |
| **5-6** | 0.29 (± 0.30) | | | | 0.31 (± 0.34) | | | | **Diagnosis x trial number + sex** | Trial number | F_3.2233_ = 9.13 | < 0.0001 |
| **7-8** | 0.26 (± 0.29) | | | | 0.28 (± 0.32) | | | |  | Diagnosis | F_1.380_ = 3.32 | 0.069 |
|  | **Pairwise differences** | | | | | | | |  | Diagnosis x trial number | F_3.2233_ = 1.96 | 0.12 |
|  | **Estimate** | **T-ratio** | **P-value** | **Adjusted p-value** | **Estimate** | **T-ratio** | **P-value** | **Adjusted p-value** |  | Sex | F_1,380_ = 6.46 | 0.01 |
| **Trials 1-2 vs. 3-4** | 0.01 | 0.30 | 0.76 | 1.00 | 0.03 | 1.82 | 0.070 | 0.070 | **Diagnosis x trial number + age** | Trial number | F_3,2233_ = 9.12 | < 0.0001 |
| **Trials 1-2 vs. 5-6** | 0.01 | 0.26 | 0.79 | 1.00 | 0.06 | 3.28 | 0.0011 | 0.0022 |  | Diagnosis | F_1,380_ = 3.26 | 0.072 |
| **Trials 1-2 vs. 7-8** | 0.04 | 1.63 | 0.100 | 0.31 | 0.10 | 5.28 | <0.0001 | 0.00027 |  | Diagnosis x trial number | F_3,2233_ = 1.95 | 0.12 |
|  | **Estimate** | | **T-ratio** | | **P-value** | | **Adjusted p-value** | |  | Age | F_1,380_ = 0.01 | 0.93 |
| **Autism trials 1-2 vs. NT 1-2** | -0.08 | | -2.65 | | 0.0085 | | 0.034 | |  | | | |
| **Autism trials 3-4 vs. NT 3-4** | -0.05 | | -1.79 | | 0.075 | | 0.22 | |  |  |  |  |
| **Autism trials 5-6 vs. NT 5-6** | -0.03 | | -0.88 | | 0.38 | | 0.76 | |  |  |  |  |
| **Autism trials 7-8 vs. NT 7-8** | -0.02 | | -0.59 | | 0.55 | | 0.76 | |  |  |  |  |

**Table S2A | Long-term habituation of the acoustically evoked eyelid startle response in children with autism compared to neurotypical children.**

All statistical comparisons done using an ANOVA on Linear Mixed-Effect Model.

P-value adjustments: Bonferroni-Holm method for three tests for within group comparisons and four tests for between group comparisons.

NEC = Normalized Eyelid Closure, SD = Standard Deviation, NT = Neurotypical.

| **Stimulus type** | **Eyelid startle amplitude regression Pearson correlation coefficient (mean NEC ± SD)** | | **Model** | **Main effect** | **B** | **SE** | **CI low** | **CI high** | **T-statistic** | **Adjusted R^2^** | **P-value** | **Adjusted p-value** |
| --- | --- | --- | --- | --- | --- | --- | --- | --- | --- | --- | --- | --- |
| **Pulse only** | **NT (n = 147)** | **Autism**  **(n = 236)** | Diagnosis | Diagnosis | -0.12 | 0.05 | -0.21 | -0.03 | -2.53 | 0.014 | 0.012 | 0.048 |
|  | -0.04 (± 0.42) | -0.16 (± 0.47) | Diagnosis + sex | Diagnosis | -0.12 | 0.05 | -0.21 | -0.02 | -2.43 | 0.012 | 0.016 | 0.062 |
|  |  |  |  | Sex | -0.02 | 0.05 | -0.12 | 0.08 | -0.43 | 0.012 | 0.67 | 1.000 |
|  |  |  | Diagnosis + age | Diagnosis | -0.14 | 0.05 | -0.23 | -0.04 | -2.75 | 0.015 | 0.006 | 0.025 |
|  |  |  |  | Age | 0.01 | 0.01 | -0.01 | 0.04 | 1.15 | 0.015 | 0.25 | 0.62 |
| **PP 5% + Pulse** | **NT (n = 147)** | **Autism**  **(n = 234)** | Diagnosis | Diagnosis | -0.05 | 0.05 | -0.14 | 0.05 | -0.98 | 0.000 | 0.33 | 0.33 |
|  | -0.12 (± 0.41) | -0.17 (± 0.49) | Diagnosis + sex | Diagnosis | -0.06 | 0.05 | -0.16 | 0.03 | -1.26 | 0.007 | 0.21 | 0.21 |
|  |  |  |  | Sex | 0.10 | 0.05 | 0.00 | 0.19 | 1.89 | 0.007 | 0.059 | 0.24 |
|  |  |  | Diagnosis + age | Diagnosis | -0.06 | 0.05 | -0.16 | 0.04 | -1.11 | -0.002 | 0.27 | 0.27 |
|  |  |  |  | Age | 0.01 | 0.01 | -0.02 | 0.03 | 0.63 | -0.002 | 0.530 | 0.62 |
| **PP 10% + Pulse** | **NT (n = 147)** | **Autism**  **(n = 236)** | Diagnosis | Diagnosis | -0.08 | 0.05 | -0.18 | 0.02 | -1.58 | 0.004 | 0.12 | 0.23 |
|  | -0.12 (± 0.46) | -0.20 (± 0.48) | Diagnosis + sex | Diagnosis | -0.08 | 0.05 | -0.18 | 0.02 | -1.63 | 0.002 | 0.10 | 0.21 |
|  |  |  |  | Sex | 0.02 | 0.05 | -0.08 | 0.12 | 0.44 | 0.002 | 0.66 | 1.000 |
|  |  |  | Diagnosis + age | Diagnosis | -0.10 | 0.05 | -0.20 | 0.00 | -1.87 | 0.006 | 0.062 | 0.12 |
|  |  |  |  | Age | 0.02 | 0.01 | -0.01 | 0.04 | 1.27 | 0.006 | 0.21 | 0.62 |
| **PP 25% + Pulse** | **NT (n = 147)** | **Autism**  **(n = 235)** | Diagnosis | Diagnosis | -0.10 | 0.05 | -0.20 | 0.00 | -1.89 | 0.007 | 0.059 | 0.18 |
|  | -0.04 (± 0.47) | -0.14 (± 0.50) | Diagnosis + sex | Diagnosis | -0.10 | 0.05 | -0.20 | 0.01 | -1.84 | 0.004 | 0.067 | 0.200 |
|  |  |  |  | Sex | -0.01 | 0.05 | -0.11 | 0.09 | -0.19 | 0.004 | 0.85 | 1.000 |
|  |  |  | Diagnosis + age | Diagnosis | -0.12 | 0.05 | -0.23 | -0.02 | -2.29 | 0.011 | 0.023 | 0.068 |
|  |  |  |  | Age | 0.02 | 0.01 | 0.00 | 0.05 | 1.67 | 0.011 | 0.096 | 0.39 |

**Table S2B | Pearson correlation coefficients for regression of the acoustically evoked eyelid startle response during PPI in autistic children compared to neurotypical children.**

All statistical comparisons done using a Linear Mixed-Effect Model.

P-value adjustments: Bonferroni-Holm method for four tests.

NEC = Normalized Eyelid Closure, SD = Standard Deviation, SE = Standard Error, CI = 95% Confidence Interval, NT = Neurotypical.

|  | **Rhythmic stimulus pattern** | | | | | | | | | **Random stimulus pattern** | | | | | | | |
| --- | --- | --- | --- | --- | --- | --- | --- | --- | --- | --- | --- | --- | --- | --- | --- | --- | --- |
|  | **Eyelid startle amplitude (mean NEC ± SD)** | | | | | | | | | **Eyelid startle amplitude (mean NEC ± SD)** | | | | | | | |
| **Pulse number** | **NT (n = 122)** | | | | **Autism (n = 213)** | | | | | **NT (n = 122)** | | | | **Autism (n = 215)** | | | |
| **1** | 0.24 (± 0.28) | | | | 0.25 (± 0.32) | | | | | 0.23 (± 0.28) | | | | 0.25 (± 0.31) | | | |
| **2** | 0.22 (± 0.30) | | | | 0.25 (± 0.34) | | | | | 0.22 (± 0.30) | | | | 0.24 (± 0.34) | | | |
| **3** | 0.25 (± 0.32) | | | | 0.26 (± 0.35) | | | | | 0.24 (± 0.31) | | | | 0.28 (± 0.35) | | | |
| **4** | 0.24 (± 0.33) | | | | 0.25 (± 0.34) | | | | | 0.20 (± 0.30) | | | | 0.23 (± 0.33) | | | |
| **5** | 0.23 (± 0.32) | | | | 0.25 (± 0.35) | | | | | 0.23 (± 0.32) | | | | 0.26 (± 0.35) | | | |
| **6** | 0.24 (± 0.30) | | | | 0.27 (± 0.33) | | | | | 0.21 (± 0.32) | | | | 0.23 (± 0.34) | | | |
|  | **Model: Diagnosis** | | | | | | | | | | | | | | | | |
| **Main effect of pulse** | F_5,16840_ = 3.25, p = 0.0062 | | | | | | | | | F_5,16767_ = 12.56, p < 0.0001 | | | | | | | |
| **Main effect of diagnosis** | F_1,332_ = 1.07, p = 0.30 | | | | | | | | | F_1,334_ = 1.07, p = 0.30 | | | | | | | |
| **Diagnosis x pulse** | F_5,16840_ = 1.25, p = 0.29 | | | | | | | | | F_5,16767_ = 0.66, p = 0.66 | | | | | | | |
|  | **Model: Diagnosis + sex** | | | | | | | | | | | | | | | | |
| **Main effect of pulse** | F_5,16840_ = 3.25, p = 0.0062 | | | | | | | | F_5,16767_ = 12.56, p < 0.0001 | | | | | | | | |
| **Main effect of diagnosis** | F_1,331_ = 1.07, p = 0.30 | | | | | | | | F_1,333_ = 1.08, p = 0.30 | | | | | | | | |
| **Main effect of sex** | F_1,331_ = 0.75, p = 0.39 | | | | | | | | F_1,333_ = 3.28, p = 0.071 | | | | | | | | |
| **Diagnosis x pulse** | F_5,16840_ = 1.25, p = 0.29 | | | | | | | | F_5,16767_ = 0.66, p = 0.66 | | | | | | | | |
|  | **Model: Diagnosis + age** | | | | | | | | | | | | | | | | |
| **Main effect of pulse** | F_5,16840_ = 3.25, p = 0.0062 | | | | | | | | F_5,16767_ = 12.56, p < 0.0001 | | | | | | | | |
| **Main effect of diagnosis** | F_1,331_ = 1.07, p = 0.30 | | | | | | | | F_1,333_ = 1.06, p = 0.30 | | | | | | | | |
| **Main effect of age** | F_1,331_ = 0.11, p = 0.74 | | | | | | | | F_1,333_ = 0.03, p = 0.87 | | | | | | | | |
| **Diagnosis x pulse** | F_5,16840_ = 1.25, p = 0.29 | | | | | | | | F_5,16767_ = 0.66, p = 0.66 | | | | | | | | |
| **Pairwise comparisons** | | | | | | | | | | | | | | | | | |
|  | **Estimate** | **T-ratio** | **P-value** | **Adjusted**  **p-value** | **Estimate** | **T-ratio** | **P-value** | **Adjusted p-value** | | **Estimate** | **T-ratio** | **P-value** | **Adjusted p-value** | **Estimate** | **T-ratio** | **P-value** | **Adjusted p-value** |
| **Pulse 1 vs. Pulse 2** | 0.02 | 2.09 | 0.037 | 0.18 | 0.01 | 0.63 | 0.53 | 1.00 | | 0.01 | 0.60 | 0.55 | 1.00 | 0.01 | 1.31 | 0.19 | 0.32 |
| **Pulse 1 vs. Pulse 3** | -0.008 | -0.75 | 0.45 | 1.00 | -0.01 | -0.81 | 0.42 | 1.00 | | -0.01 | -1.14 | 0.25 | 0.76 | -0.03 | -3.19 | 0.0014 | 0.0070 |
| **Pulse 1 vs. Pulse 4** | 0.005 | 0.47 | 0.64 | 1.00 | 0.004 | 0.45 | 0.66 | 1.00 | | 0.03 | 2.51 | 0.012 | 0.061 | 0.02 | 2.12 | 0.034 | 0.10 |
| **Pulse 1 vs. Pulse 5** | 0.01 | 0.96 | 0.34 | 1.00 | 0.01 | 0.69 | 0.49 | 1.00 | | -0.001 | -0.13 | 0.90 | 1.00 | -0.01 | -1.41 | 0.16 | 0.32 |
| **Pulse 1 vs. Pulse 6** | 0.006 | 0.53 | 0.60 | 1.00 | -0.02 | -2.29 | 0.022 | 0.11 | | 0.02 | 1.82 | 0.069 | 0.28 | 0.02 | 2.43 | 0.015 | 0.060 |
| **NT vs. autism** | **Estimate** | | **T-ratio** | | **P-value** | | **Adjusted p-value** | | | **Estimate** | | **T-ratio** | | **P-value** | | **Adjusted p-value** | |
| **Pulse 1 vs. Pulse 1** | -0.02 | | -0.64 | | 0.53 | | 1.00 | | | -0.02 | | -0.75 | | 0.45 | | 1.00 | |
| **Pulse 2 vs. Pulse 2** | -0.03 | | -1.35 | | 0.18 | | 0.90 | | | -0.01 | | -0.54 | | 0.59 | | 1.00 | |
| **Pulse 3 vs. Pulse 3** | -0.01 | | -0.59 | | 0.56 | | 1.00 | | | -0.03 | | -1.40 | | 0.16 | | 0.97 | |
| **Pulse 4 vs. Pulse 4** | -0.02 | | -0.69 | | 0.49 | | 1.00 | | | -0.03 | | -1.13 | | 0.26 | | 1.00 | |
| **Pulse 5 vs. Pulse 5** | -0.02 | | -0.82 | | 0.41 | | 1.00 | | | -0.03 | | -1.21 | | 0.23 | | 1.00 | |
| **Pulse 6 vs. Pulse 6** | -0.04 | | -1.69 | | 0.092 | | 0.55 | | | -0.02 | | -0.70 | | 0.49 | | 1.00 | |

**Table S3A | Short-term habituation of the acoustically evoked eyelid startle response using rhythmic and random stimulus patterns in children with autism compared to neurotypical children.**

All statistical comparisons done using an ANOVA on Linear Mixed-Effect Model.

P-value adjustment: Bonferroni-Holm method for five tests for within group comparisons and six tests for between group comparisons.

NEC = Normalized Eyelid Closure, SD = standard deviation, NT = Neurotypical.

| **Stimulus type** | **Eyelid closure amplitude (mean NEC ± SD)** | | **Model** | **Main effect** | **B** | **SE** | **CI low** | **CI high** | **T-statistic** | **Adjusted R^2^** | **P-value** | **Adjusted p-value** |
| --- | --- | --- | --- | --- | --- | --- | --- | --- | --- | --- | --- | --- |
| **Rhythmic pattern observation window 1** | **NT (n = 122)** | **Autism (n = 213)** | Diagnosis | Diagnosis | 0.01 | 0.02 | -0.03 | 0.05 | 0.58 | 0.000 | 0.56 | - |
|  | 0.16 (± 0.31) | 0.17 (± 0.31) | Diagnosis + sex | Diagnosis | 0.02 | 0.020 | -0.02 | 0.06 | 0.84 | 0.004 | 0.40 |  |
|  |  |  |  | Sex | -0.04 | 0.02 | -0.08 | 0.00 | -1.88 |  | 0.06 |  |
|  |  |  | Diagnosis + age | Diagnosis | 0.00 | 0.02 | -0.04 | 0.04 | 0.03 | 0.005 | 0.97 |  |
|  |  |  |  | Age | 0.01 | 0.00 | 0.00 | 0.02 | 2.32 |  | 0.02 |  |
| **Random pattern observation window 1** | **NT (n = 122)** | **Autism (n = 215)** | Diagnosis | Diagnosis | 0.02 | 0.02 | -0.02 | 0.07 | 1.13 | 0.001 | 0.26 | 0.26 |
|  | 0.15 (± 0.30) | 0.17 (± 0.33) | Diagnosis + sex | Diagnosis | 0.03 | 0.02 | -0.01 | 0.07 | 1.38 | 0.005 | 0.17 | 0.17 |
|  |  |  |  | Sex | -0.04 | 0.02 | -0.08 | 0.00 | -1.83 |  | 0.07 | 0.14 |
|  |  |  | Diagnosis + age | Diagnosis | 0.01 | 0.02 | -0.03 | 0.06 | 0.61 | 0.006 | 0.54 | 0.54 |
|  |  |  |  | Age | 0.01 | 0.01 | 0.00 | 0.02 | 2.19 |  | 0.029 | 0.029 |
| **Random pattern observation window 2** | **NT (n = 122)** | **Autism (n = 215)** | Diagnosis | Diagnosis | 0.04 | 0.02 | 0.00 | 0.07 | 2.03 | 0.003 | 0.04 | 0.09 |
|  | 0.13 (± 0.29) | 0.17 (± 0.32) | Diagnosis + sex | Diagnosis | 0.04 | 0.02 | 0.00 | 0.07 | 2.18 | 0.004 | 0.03 | 0.06 |
|  |  |  |  | Sex | -0.02 | 0.02 | -0.06 | 0.01 | -1.17 |  | 0.24 | 0.24 |
|  |  |  | Diagnosis + age | Diagnosis | 0.02 | 0.02 | -0.01 | 0.06 | 1.21 | 0.012 | 0.23 | 0.45 |
|  |  |  |  | Age | 0.01 | 0.00 | 0.01 | 0.02 | 3.46 |  | 0.001 | 0.001 |

**Table S3B | Amplitude of anticipatory eyeblinks in predefined observation windows for rhythmic and random stimulus patterns in autistic compared to neurotypical children.**

All statistical comparisons done using Linear Mixed-Effect Models.

P-value adjustment: Bonferroni-Holm method for two tests.

NEC = Normalized Eyelid Closure, SD = Standard Deviation, SE = Standard Error, CI = 95% confidence interval, NT = Neurotypical.

| **Stimulus Pattern** | **Cumulative sum of eyelid amplitude (mean NEC ± SD)** | | **Model** | **Main effect** | **B** | **SE** | **CI low** | **CI high** | **T-statistic** | **Adjusted R^2^** | **P-value** |
| --- | --- | --- | --- | --- | --- | --- | --- | --- | --- | --- | --- |
| **Rhythmic** | **NT (n = 122)** | **Autism (n = 213)** | Diagnosis | Diagnosis | 18.06 | 6.14 | 5.97 | 30.15 | 2.94 | 0.01 | 0.0035 |
|  | 61.00 (± 75.42) | 78.36 (± 108.98) | Diagnosis + sex | Diagnosis | 18.67 | 6.21 | 6.45 | 30.89 | 3.01 | 0.01 | 0.003 |
|  |  |  |  | Sex | -4.54 | 6.39 | -17.10 | 8.03 | -0.71 | 0.01 | 0.48 |
|  |  |  | Diagnosis + age | Diagnosis | 15.40 | 6.29 | 3.03 | 27.76 | 2.45 | 0.01 | 0.015 |
|  |  |  |  | Age | 2.72 | 1.48 | -0.19 | 5.62 | 1.84 | 0.01 | 0.067 |
|  | **SD of max cumulative sum** | |  |  |  |  |  |  |  |  |  |
|  | 54.22 (± 33.52) | 87.14 (± 48.96) | Diagnosis | Diagnosis | 32.91 | 4.99 | 23.10 | 42.73 | 6.60 | 0.11 | <0.0001 |
|  |  |  | Diagnosis + sex | Diagnosis | 32.37 | 5.04 | 22.46 | 42.28 | 6.43 | 0.11 | <0.0001 |
|  |  |  |  | Sex | 4.14 | 5.16 | -6.01 | 14.30 | 0.80 | 0.11 | 0.42 |
|  |  |  | Diagnosis + age | Diagnosis | 30.61 | 5.10 | 20.58 | 40.64 | 6.00 | 0.12 | <0.0001 |
|  |  |  |  | Age | 2.38 | 1.19 | 0.04 | 4.73 | 2.00 | 0.12 | 0.047 |
| **Random** | **Cumulative sum of eyelid amplitude (mean NEC ± SD)** | |  |  |  |  |  |  |  |  |  |
|  | **NT (n = 122)** | **Autism (n = 215)** | Diagnosis | Diagnosis | 19.16 | 6.41 | 6.54 | 31.78 | 2.99 | 0.01 | 0.003 |
|  | 63.92 (± 83.17) | 81.90 (± 114.52) | Diagnosis + sex | Diagnosis | 20.26 | 6.47 | 7.53 | 33.00 | 3.13 | 0.01 | 0.002 |
|  |  |  |  | Sex | -8.21 | 6.66 | -21.32 | 4.90 | -1.23 | 0.01 | 0.22 |
|  |  |  | Diagnosis + age | Diagnosis | 15.48 | 6.54 | 2.62 | 28.35 | 2.37 | 0.01 | 0.019 |
|  |  |  |  | Age | 3.73 | 1.53 | 0.71 | 6.75 | 2.43 | 0.01 | 0.016 |
|  | **SD of max cumulative sum** | |  |  |  |  |  |  |  |  |  |
|  | 60.64 (± 37.30) | 89.55 (± 52.52) | Diagnosis | Diagnosis | 28.91 | 5.39 | 18.30 | 39.52 | 5.36 | 0.08 | <0.0001 |
|  |  |  | Diagnosis + sex | Diagnosis | 28.74 | 5.45 | 18.02 | 39.46 | 5.27 | 0.07 | <0.0001 |
|  |  |  |  | Sex | 1.25 | 5.59 | -9.74 | 12.23 | 0.22 | 0.07 | 0.82 |
|  |  |  | Diagnosis + age | Diagnosis | 28.09 | 5.54 | 17.19 | 38.99 | 5.07 | 0.07 | <0.0001 |
|  |  |  |  | Age | 0.84 | 1.30 | -1.71 | 3.39 | 0.65 | 0.07 | 0.52 |

**Table S3C | Maximum cumulative sum of eyelid closure amplitude and the associated variability for rhythmic and random stimulus patterns in children with autism compared to neurotypical children.**

All statistical comparisons done using Linear Mixed-Effect Models.

NEC = Normalized Eyelid Closure, SD = Standard Deviation, SE = Standard Error, CI = 95% confidence interval, NT = Neurotypical.

| **Behavioral response** | **Mean ± SD** | | **Model** | **Main effect** | **B** | **SE** | **OR** | **CI low** | **CI high** | **T-statistic** | **Adjusted R^2^** | **P-value** | **Adjusted p-value** |
| --- | --- | --- | --- | --- | --- | --- | --- | --- | --- | --- | --- | --- | --- |
|  | **NT (n = 156)** | **Autism (n = 275)** |  |  |  |  |  |  |  |  |  |  |  |
| **Screen avoidance (% of frames)** | 3.66 (± 7.76) | 9.94 (± 11.56) | Diagnosis | Diagnosis | 1.81 | 0.20 | 6.09 | 4.16 | 8.93 | 9.26 | 0.11 | <0.0001 | <0.0001 |
|  |  |  | Diagnosis + sex | Diagnosis | 1.80 | 0.20 | 6.02 | 4.10 | 8.85 | 9.13 | 0.11 | <0.0001 | <0.0001 |
|  |  |  |  | Sex | 0.08 | 0.19 | 1.09 | 0.75 | 1.57 | 0.45 | 0.11 | 0.65 | 1.00 |
|  |  |  | Diagnosis + age | Diagnosis | 1.98 | 0.20 | 7.26 | 4.91 | 10.75 | 9.91 | 0.13 | <0.0001 | <0.0001 |
|  |  |  |  | Age | -0.17 | 0.04 | 0.84 | 0.77 | 0.92 | -3.86 | 0.13 | 0.00 | 0.001 |
| **Headphone touches (% of trials)** | 1.87 (± 5.97) | 11.31 (± 16.74) | Diagnosis | Diagnosis | 2.81 | 0.25 | 16.67 | 10.28 | 27.04 | 11.40 | 0.20 | <0.0001 | <0.0001 |
|  |  |  | Diagnosis + sex | Diagnosis | 2.78 | 0.25 | 16.09 | 9.90 | 26.15 | 11.21 | 0.20 | <0.0001 | <0.0001 |
|  |  |  |  | Sex | 0.30 | 0.23 | 1.36 | 0.87 | 2.12 | 1.34 | 0.20 | 0.18 | 1.00 |
|  |  |  | Diagnosis + age | Diagnosis | 2.94 | 0.26 | 18.84 | 11.38 | 31.16 | 11.43 | 0.21 | <0.0001 | <0.0001 |
|  |  |  |  | Age | -0.11 | 0.05 | 0.90 | 0.81 | 1.00 | -1.95 | 0.21 | 0.05 | 0.36 |
| **Non-syllabic vocalizations (% of trials)** | 0.43 (± 2.40) | 11.16 (± 15.21) | Diagnosis | Diagnosis | 4.56 | 0.38 | 95.33 | 45.44 | 199.97 | 12.06 | 0.39 | <0.0001 | <0.0001 |
|  |  |  | Diagnosis + sex | Diagnosis | 4.61 | 0.38 | 100.87 | 48.27 | 210.76 | 12.27 | 0.40 | <0.0001 | <0.0001 |
|  |  |  |  | Sex | -0.52 | 0.27 | 0.60 | 0.35 | 1.01 | -1.91 | 0.40 | 0.06 | 0.45 |
|  |  |  | Diagnosis + age | Diagnosis | 4.50 | 0.38 | 90.12 | 42.50 | 191.09 | 11.74 | 0.39 | <0.0001 | <0.0001 |
|  |  |  |  | Age | 0.04 | 0.07 | 1.04 | 0.92 | 1.19 | 0.67 | 0.39 | 0.50 | 1.00 |
|  |  |  |  |  |  |  | **β** |  |  |  |  |  |  |
| **AP postural stability (mean SD over all trials)** | 2.11 (± 1.77) | 4.24 (± 2.82) | Diagnosis | Diagnosis | 1.00 | 0.09 | 0.48 | 0.83 | 1.17 | 11.36 | 0.23 | <0.0001 | <0.0001 |
|  |  |  | Diagnosis + sex | Diagnosis | 0.99 | 0.09 | 0.47 | 0.81 | 1.16 | 11.06 | 0.23 | <0.0001 | <0.0001 |
|  |  |  |  | Sex | 0.09 | 0.09 |  | -0.09 | 0.27 | 1.01 | 0.23 | 0.31 | 1.00 |
|  |  |  | Diagnosis + age | Diagnosis | 1.06 | 0.09 | 0.51 | 0.88 | 1.23 | 11.69 | 0.24 | <0.0001 | <0.0001 |
|  |  |  |  | Age | -0.05 | 0.02 |  | -0.10 | -0.01 | -2.49 | 0.24 | 0.01 | 0.12 |
| **Head rotations (% of trials)** | 16.62 (± 16.76) | 40.22 (± 25.62) | Diagnosis | Diagnosis | 1.12 | 0.08 | 0.54 | 0.95 | 1.28 | 13.21 | 0.29 | <0.0001 | <0.0001 |
|  |  |  | Diagnosis + sex | Diagnosis | 1.11 | 0.09 | 0.53 | 0.94 | 1.28 | 12.96 | 0.29 | <0.0001 | <0.0001 |
|  |  |  |  | Sex | 0.04 | 0.09 |  | -0.13 | 0.21 | 0.49 | 0.29 | 0.63 | 1.00 |
|  |  |  | Diagnosis + age | Diagnosis | 1.12 | 0.09 | 0.54 | 0.94 | 1.29 | 12.73 | 0.29 | <0.0001 | <0.0001 |
|  |  |  |  | Age | 0.00 | 0.02 |  | -0.04 | 0.04 | 0.11 | 0.29 | 0.92 | 1.00 |
| **Mouth openings (mean area over all trials)** | 2.96 (± 3.02) | 9.55 (± 6.95) | Diagnosis | Diagnosis | 1.13 | 0.08 | 0.55 | 0.97 | 1.30 | 13.47 | 0.30 | <0.0001 | <0.0001 |
|  |  |  | Diagnosis + sex | Diagnosis | 1.12 | 0.09 | 0.54 | 0.95 | 1.29 | 13.14 | 0.30 | <0.0001 | <0.0001 |
|  |  |  |  | Sex | 0.09 | 0.09 |  | -0.08 | 0.26 | 1.08 | 0.30 | 0.28 | 1.00 |
|  |  |  | Diagnosis + age | Diagnosis | 1.12 | 0.09 | 0.54 | 0.95 | 1.29 | 12.84 | 0.29 | <0.0001 | <0.0001 |
|  |  |  |  | Age | 0.01 | 0.02 |  | -0.03 | 0.05 | 0.71 | 0.29 | 0.48 | 1.00 |
| **Pupil range (mean horizontal movement over all trials)** | 10.12 (± 3.48) | 14.33 (± 4.04) | Diagnosis | Diagnosis | 4.21 | 0.39 | 0.47 | 3.45 | 4.97 | 10.92 | 0.22 | <0.0001 | <0.0001 |
|  |  |  | Diagnosis + sex | Diagnosis | 4.14 | 0.39 | 0.46 | 3.38 | 4.91 | 10.62 | 0.22 | <0.0001 | <0.0001 |
|  |  |  |  | Sex | 0.42 | 0.40 |  | -0.35 | 1.20 | 1.07 | 0.22 | 0.29 | 1.00 |
|  |  |  | Diagnosis + age | Diagnosis | 4.23 | 0.40 | 0.47 | 3.44 | 5.01 | 10.59 | 0.21 | <0.0001 | <0.0001 |
|  |  |  |  | Age | -0.01 | 0.09 |  | -0.20 | 0.17 | -0.16 | 0.21 | 0.88 | 1.00 |
| **Side eye** | 0.09 (± 0.08) | 0.13 (± 0.08) | Diagnosis | Diagnosis | 0.55 | 0.10 | 0.27 | 0.36 | 0.74 | 5.73 | 0.07 | <0.0001 | <0.0001 |
|  |  |  | Diagnosis + sex | Diagnosis | 0.53 | 0.10 | 0.26 | 0.34 | 0.72 | 5.43 | 0.07 | <0.0001 | <0.0001 |
|  |  |  |  | Sex | 0.16 | 0.10 |  | -0.04 | 0.35 | 1.58 | 0.07 | 0.12 | 0.81 |
|  |  |  | Diagnosis + age | Diagnosis | 0.54 | 0.10 | 0.26 | 0.35 | 0.74 | 5.42 | 0.07 | <0.0001 | <0.0001 |
|  |  |  |  | Age | 0.01 | 0.02 |  | -0.04 | 0.06 | 0.46 | 0.07 | 0.65 | 1.00 |
| **Baseline regression intercept** | 4.77 (± 3.51) | 8.31 (± 5.55) | Diagnosis | Diagnosis | 0.75 | 0.09 | 0.36 | 0.57 | 0.94 | 8.04 | 0.13 | <0.0001 | <0.0001 |
|  |  |  | Diagnosis + sex | Diagnosis | 0.72 | 0.09 | 0.35 | 0.54 | 0.91 | 7.65 | 0.13 | <0.0001 | <0.0001 |
|  |  |  |  | Sex | 0.19 | 0.10 |  | 0.00 | 0.38 | 2.00 | 0.13 | 0.05 | 0.41 |
|  |  |  | Diagnosis + age | Diagnosis | 0.73 | 0.10 | 0.35 | 0.54 | 0.92 | 7.59 | 0.13 | <0.0001 | <0.0001 |
|  |  |  |  | Age | 0.02 | 0.02 |  | -0.03 | 0.06 | 0.71 | 0.13 | 0.48 | 1.00 |
| **Baseline regression SD** | 3.27 (± 1.85) | 5.23 (± 2.60) | Diagnosis | Diagnosis | 0.83 | 0.09 | 0.40 | 0.65 | 1.02 | 9.08 | 0.16 | <0.0001 | <0.0001 |
|  |  |  | Diagnosis + sex | Diagnosis | 0.80 | 0.09 | 0.39 | 0.62 | 0.99 | 8.67 | 0.17 | <0.0001 | <0.0001 |
|  |  |  |  | Sex | 0.20 | 0.09 |  | 0.01 | 0.38 | 2.12 | 0.17 | 0.03 | 0.35 |
|  |  |  | Diagnosis + age | Diagnosis | 0.78 | 0.09 | 0.38 | 0.60 | 0.97 | 8.26 | 0.17 | <0.0001 | <0.0001 |
|  |  |  |  | Age | 0.05 | 0.02 |  | 0.01 | 0.09 | 2.20 | 0.17 | 0.03 | 0.23 |

**Table S4 | Smartphone-based assessments of behavioral responses in autistic children compared to neurotypical children.**

Multilevel binomial logistic regression for screen avoidance, headphone touches, non-syllabic vocalizations.

Linear regression for: AP postural stability (log transformed); head rotations (log transformed); mouth openings (square root transformed); side eye (square root transformed); baseline regression intercept (square root transformed); baseline regression SD (square root transformed).

P-value adjustments: Bonferroni-Holm method for 10 tests.

NT = Neurotypical, SD = Standard Deviation, SE = Standard Error, CI = 95% confidence interval, OR = Odds Ratio, AP = Anteroposterior.

|  | **None** | **ADHD only** | **ID only** | **ADHD and ID** |
| --- | --- | --- | --- | --- |
| **Number of participants (%)** | 100 (36.36) | 34 (12.36) | 69 (25.09) | 50 (18.18) |
| **Mean age (SD)** | 7.97 (± 1.92) | 7.48 (± 1.98) | 8.54 (± 1.88) | 7.82 (± 1.74) |
| **% girls** | 26 | 32.35 | 28.99 | 26 |
| **% boys** | 74 | 67.65 | 71.01 | 74 |

**Table S5 | Most common co-occurring conditions in the sample of children with autism.**

ADHD = Attention Deficit Hyperactivity Disorder, ID = Intellectual Disability, SD = Standard Deviation.

| **Stimulus type** | **Autism only (n = 87)** | **ADHD**  **(n = 27)** | **ID**  **(n = 63)** | **ADHD & ID (n = 41)** | **Main effect** | **B** | **SE** | **CI low** | **CI high** | **T-statistic** | **Adjusted R^2^** | **P-value** | **Adjusted p-value** |
| --- | --- | --- | --- | --- | --- | --- | --- | --- | --- | --- | --- | --- | --- |
|  | **Eyelid startle amplitude (mean NEC ± SD)** | | | |  |  |  |  |  |  |  |  |  |
| **Pulse only** | 0.39 (± 0.35) | 0.35 (± 0.34) | 0.33 (± 0.34) | 0.28 (± 0.32) | ADHD | -0.02 | 0.05 | -0.13 | 0.08 | -0.46 | 0.010 | 0.65 | - |
|  |  |  |  |  | ID | -0.05 | 0.04 | -0.13 | 0.03 | -1.32 | 0.010 | 0.19 | - |
|  |  |  |  |  | ADHD & ID | -0.10 | 0.05 | -0.19 | -0.01 | -2.15 | 0.010 | 0.033 | - |
| **PP 5% + Pulse** | 0.36 (± 0.34) | 0.32 (± 0.35) | 0.32 (± 0.36) | 0.29 (± 0.33) | ADHD | -0.04 | 0.05 | -0.15 | 0.06 | -0.79 | 0.005 | 0.43 | 0.81 |
|  |  |  |  |  | ID | -0.05 | 0.04 | -0.13 | 0.03 | -1.21 | 0.005 | 0.23 | 0.23 |
|  |  |  |  |  | ADHD & ID | -0.08 | 0.05 | -0.18 | 0.01 | -1.68 | 0.005 | 0.095 | 0.29 |
| **PP 10% + Pulse** | 0.36 (± 0.37) | 0.32 (± 0.36) | 0.28 (± 0.34) | 0.33 (± 0.35) | ADHD | -0.04 | 0.05 | -0.15 | 0.06 | -0.84 | 0.009 | 0.40 | 0.81 |
|  |  |  |  |  | ID | -0.09 | 0.04 | -0.17 | -0.01 | -2.22 | 0.009 | 0.027 | 0.055 |
|  |  |  |  |  | ADHD & ID | -0.02 | 0.05 | -0.11 | 0.07 | -0.40 | 0.009 | 0.69 | 0.69 |
| **PP 25% + Pulse** | 0.36 (± 0.35) | 0.28 (± 0.30) | 0.25 (± 0.33) | 0.31 (± 0.34) | ADHD | -0.08 | 0.05 | -0.18 | 0.02 | -1.57 | 0.014 | 0.12 | 0.35 |
|  |  |  |  |  | ID | -0.12 | 0.04 | -0.19 | -0.04 | -3.04 | 0.014 | 0.003 | **0.008** |
|  |  |  |  |  | ADHD & ID | -0.05 | 0.04 | -0.14 | 0.04 | -1.13 | 0.014 | 0.26 | 0.52 |
|  | **Variability in eyelid startle amplitude (mean NEC ± SD)** | | | | |  |  |  |  |  |  |  |  |
| **Pulse only** | 0.24 (± 0.12) | 0.22 (± 0.11) | 0.26 (± 0.12) | 0.27 (± 0.10) | ADHD | -0.02 | 0.03 | -0.07 | 0.03 | -0.86 | 0.002 | 0.39 | - |
|  |  |  |  |  | ID | 0.02 | 0.02 | -0.02 | 0.05 | 0.79 | 0.002 | 0.43 | - |
|  |  |  |  |  | ADHD & ID | 0.03 | 0.02 | -0.02 | 0.07 | 1.20 | 0.002 | 0.23 | - |
| **PP 5% + Pulse** | 0.23 (± 0.10) | 0.24 (± 0.12) | 0.26 (± 0.14) | 0.26 (± 0.12) | ADHD | 0.01 | 0.03 | -0.04 | 0.06 | 0.29 | -0.004 | 0.77 | 0.77 |
|  |  |  |  |  | ID | 0.03 | 0.02 | -0.01 | 0.07 | 1.36 | -0.004 | 0.18 | 0.35 |
|  |  |  |  |  | ADHD & ID | 0.02 | 0.02 | -0.02 | 0.07 | 0.98 | -0.004 | 0.33 | 0.33 |
| **PP 10% + Pulse** | 0.26 (± 0.12) | 0.29 (± 0.15) | 0.23 (± 0.11) | 0.30 (± 0.12) | ADHD | 0.03 | 0.03 | -0.02 | 0.08 | 1.20 | 0.031 | 0.23 | 0.46 |
|  |  |  |  |  | ID | -0.03 | 0.02 | -0.07 | 0.01 | -1.57 | 0.031 | 0.12 | 0.35 |
|  |  |  |  |  | ADHD & ID | 0.04 | 0.02 | -0.01 | 0.08 | 1.63 | 0.031 | 0.10 | 0.31 |
| **PP 25% + Pulse** | 0.24 (± 0.11) | 0.20 (± 0.10) | 0.25 (± 0.12) | 0.28 (± 0.09) | ADHD | -0.05 | 0.02 | -0.10 | 0.00 | -1.99 | 0.027 | 0.048 | 0.15 |
|  |  |  |  |  | ID | 0.01 | 0.02 | -0.03 | 0.05 | 0.51 | 0.027 | 0.61 | 0.61 |
|  |  |  |  |  | ADHD & ID | 0.03 | 0.02 | -0.01 | 0.07 | 1.50 | 0.027 | 0.14 | 0.31 |

**Table S6A | Prepulse inhibition of the acoustically evoked eyelid startle response in autistic children with and without co-occurring conditions.**

All statistical comparisons done using Linear Mixed-Effect Models.

P-value adjustments: Bonferroni-Holm method for three tests for eyelid startle amplitude comparisons and for four tests for variability in eyelid startle amplitude comparisons.

ADHD = Attention Deficit Hyperactivity Disorder, ID = Intellectual Disability, SE = Standard Error, CI = 95% confidence interval, NEC = Normalized Eyelid Closure. SD = Standard Deviation, PP = prepulse.

| **ANOVA on LME for eyelid startle amplitude** | | | | |
| --- | --- | --- | --- | --- |
|  | | | **Main effect trial type** | |
|  |  |  | **F-test** | **P-value** |
| **None (n = 87)** | | | F_3,2049_ = 1.94 | 0.12 |
| **ADHD (n = 27)** | | | F_3,717_ = 2.66 | 0.047 |
| **ID (n = 63)** | | | F_3.1623_ = 8.73 | <0.0001 |
| **ADHD & ID (n = 41)** | | | F_3,1013_ = 1.86 | 0.13 |
| **Pairwise differences** | | | | |
|  | **Estimate** | **T-ratio** | **P-value** | **Adjusted**  **p-value** |
| **None** |  | | | |
| **(Pulse) vs. (PP 5% + Pulse)** | 0.03 | 1.57 | 0.14 | 0.14 |
| **(Pulse) vs. (PP 10% + Pulse)** | 0.03 | 1.87 | 0.061 | 0.12 |
| **(Pulse) vs. (PP 25% + Pulse)** | 0.04 | 2.24 | 0.025 | 0.074 |
| **ADHD only** |  | | | |
| **(Pulse) vs. (PP 5% + Pulse)** | 0.04 | 1.32 | 0.19 | 0.34 |
| **(Pulse) vs. (PP 10% + Pulse)** | 0.04 | 1.37 | 0.17 | 0.34 |
| **(Pulse) vs. (PP 25% + Pulse)** | 0.08 | 2.82 | 0.0049 | 0.015 |
| **ID only** |  |  |  |  |
| **(Pulse) vs. (PP 5% + Pulse)** | 0.03 | 1.34 | 0.18 | 0.18 |
| **(Pulse) vs. (PP 10% + Pulse)** | 0.06 | 3.25 | 0.0012 | 0.0023 |
| **(Pulse) vs. (PP 25% + Pulse)** | 0.09 | 4.75 | <0.0001 | <0.0001 |
| **ADHD & ID** |  |  |  |  |
| **(Pulse) vs. (PP 5% + Pulse)** | 0.0009 | 0.04 | 0.97 | 0.97 |
| **(Pulse) vs. (PP 10% + Pulse)** | -0.05 | -2.00 | 0.045 | 0.14 |
| **(Pulse) vs. (PP 25% + Pulse)** | -0.02 | -0.94 | 0.35 | 0.70 |

**Table S6B | Within group effect of trial type for prepulse inhibition of the acoustically evoked eyelid startle response in children with autism with and without co-occurring conditions.**

All statistical comparisons done using ANOVA on Linear Mixed-Effect Model.

P-value adjustments: Bonferroni-Holm method for three tests.

LME = Linear Mixed-Effect Model, ADHD = Attention Deficit Hyperactivity Disorder, ID = Intellectual Disability, NEC = Normalized Eyelid Closure, PP = prepulse.

|  | **Eyelid startle amplitude (mean NEC ± SD)** | | | | | | | | | | | | | | | |
| --- | --- | --- | --- | --- | --- | --- | --- | --- | --- | --- | --- | --- | --- | --- | --- | --- |
| **Trial number** | **None (n = 88)** | | | | **ADHD (n = 28)** | | | | **ID (n = 64)** | | | | **ADHD & ID ( n = 42)** | | | |
| **1-2** | 0.42 (± 0.37) | | | | 0.39 (± 0.37) | | | | 0.39 (± 0.36) | | | | 0.34 (± 0.33) | | | |
| **3-4** | 0.41 (± 0.35) | | | | 0.37 (± 0.32) | | | | 0.31 (± 0.36) | | | | 0.32 (± 0.35) | | | |
| **5-6** | 0.37 (± 0.36) | | | | 0.31 (± 0.36) | | | | 0.33 (± 0.33) | | | | 0.27 (± 0.28) | | | |
| **7-8** | 0.33 (± 0.33) | | | | 0.32 (± 0.31) | | | | 0.30 (± 0.31) | | | | 0.18 (± 0.30) | | | |
| **Main effect** | **Trial number:** F_3,1192_ = 436.14, p < 0.0001 | | | | | | | | | | | | | | | |
| **Main effect** | **Co-occurring condition:** F_3,214_ = 1.66, p = 0.18 | | | | | | | | | | | | | | | |
| **Interaction effect** | **Co-occurring condition x trial number:** F_9,1192_ = 1.01, p=0.43 | | | | | | | | | | | | | | | |
|  | **Pairwise differences** | | | | | | | | | | | | | | | |
|  | **Estimate** | **T-ratio** | **P-value** | **Adjusted p-value** | **Estimate** | **T-ratio** | **P-value** | **Adjusted p-value** | **Estimate** | **T-ratio** | **P-value** | **Adjusted p-value** | **Estimate** | **T-ratio** | **P-value** | **Adjusted p-value** |
| **Trials 1-2 vs. 3-4** | 0.01 | 0.37 | 0.72 | 0.72 | 0.01 | 0.22 | 0.82 | 0.83 | 0.10 | 2.83 | 0.0047 | 0.014 | -0.02 | -0.43 | 0.67 | 0.67 |
| **Trials 1-2 vs. 5-6** | 0.04 | 1.23 | 0.22 | 0.44 | 0.07 | 1.31 | 0.19 | 0.57 | 0.07 | 1.86 | 0.063 | 0.063 | 0.05 | 1.09 | 0.27 | 0.55 |
| **Trials 1-2 vs. 7-8** | 0.10 | 2.83 | 0.0047 | 0.014 | 0.07 | 1.25 | 0.21 | 0.57 | 0.10 | 2.75 | 0.0061 | 0.014 | 0.12 | 2.52 | 0.012 | 0.035 |
|  | **Estimate** | | | | **T-ratio** | | | | **P-value** | | | | **Adjusted p-value** | | | |
| **ADHD trials 1-2 vs. none 1-2** | -0.02 | | | | -0.34 | | | | 0.73 | | | | 1.00 | | | |
| **ADHD trials 3-4 vs. none 3-4** | -0.02 | | | | -0.34 | | | | 0.74 | | | | 1.00 | | | |
| **ADHD trials 5-6 vs. NT 5-6** | -0.05 | | | | -0.80 | | | | 0.43 | | | | 1.00 | | | |
| **ADHD trials 7-8 vs. NT 7-8** | 0.01 | | | | 0.12 | | | | 0.90 | | | | 1.00 | | | |
| **ID trials 1-2 vs. none 1-2** | -0.02 | | | | -0.40 | | | | 0.69 | | | | 1.00 | | | |
| **ID trials 3-4 vs. none 3-4** | -0.11 | | | | -2.22 | | | | 0.027 | | | | 0.11 | | | |
| **ID trials 5-6 vs. none 5-6** | -0.05 | | | | -0.97 | | | | 0.34 | | | | 1.00 | | | |
| **ID trials 7-8 vs. none 7-8** | -0.03 | | | | -0.51 | | | | 0.61 | | | | 1.00 | | | |
| **Both trials 1-2 vs. none 1-2** | -0.10 | | | | -1.71 | | | | 0.089 | | | | 0.18 | | | |
| **Both trials 3-4 vs. none 3-4** | -0.07 | | | | -1.18 | | | | 0.24 | | | | 0.24 | | | |
| **Both trials 5-6 vs. none 5-6** | -0.11 | | | | -1.91 | | | | 0.058 | | | | 0.17 | | | |
| **Both trials 7-8 vs. none 7-8** | -0.12 | | | | -2.08 | | | | 0.039 | | | | 0.16 | | | |

**Table S7 | Long-term habituation of the acoustically evoked eyelid startle response in autistic children with and without co-occurring conditions.**

All statistical comparisons done using an ANOVA on Linear Mixed-Effect Model.

P-value adjustments: Bonferroni-Holm method for three tests for within group comparisons and four tests for between group comparisons.

NEC = Normalized Eyelid Closure, SD = Standard Deviation, ADHD = Attention Deficit Hyperactivity Disorder, ID = Intellectual Disability.

|  | **Rhythmic stimulus pattern** | | | | | | | | | | | | | | | |
| --- | --- | --- | --- | --- | --- | --- | --- | --- | --- | --- | --- | --- | --- | --- | --- | --- |
|  | **Eyelid startle amplitude (mean NEC ± SD)** | | | | | | | | | | | | | | | |
| **Pulse number** | **Autism only (n = 81)** | | | | **ADHD (n = 29)** | | | | **ID (n = 56)** | | | | **ADHD & ID (n = 30)** | | | |
| **1** | 0.28 (± 0.31) | | | | 0.27 (± 0.33) | | | | 0.23 (± 0.30) | | | | 0.25 (± 0.32) | | | |
| **2** | 0.26 (± 0.34) | | | | 0.24 (± 0.35) | | | | 0.22 (± 0.32) | | | | 0.27 (± 0.36) | | | |
| **3** | 0.30 (± 0.35) | | | | 0.30 (± 0.37) | | | | 0.22 (± 0.33) | | | | 0.34 (± 0.37) | | | |
| **4** | 0.24 (± 0.33) | | | | 0.25 (± 0.35) | | | | 0.20 (± 0.32) | | | | 0.25 (± 0.36) | | | |
| **5** | 0.28 (± 0.35) | | | | 0.27 (± 0.38) | | | | 0.23 (± 0.35) | | | | 0.32 (± 0.35) | | | |
| **6** | 0.24 (± 0.33) | | | | 0.23 (± 0.37) | | | | 0.20 (± 0.33) | | | | 0.27 (± 0.35) | | | |
| **Main effect** | Pulse: F_3,9463_ = 2.46, p = 0.031 | | | | | | | | | | | | | | | |
| **Main effect** | Co-occurring condition: F_3,191_ = 0.61, p = 0.61 | | | | | | | | | | | | | | | |
| **Interaction** | Pulse x co-occurring condition: F_15,9463_ = 1.17, p = 0.29 | | | | | | | | | | | | | | | |
|  | **Pairwise comparisons** | | | | | | | | | | | | | | | |
|  | **Estimate** | **T-ratio** | **P-value** | **Adjusted p-value** | **Estimate** | **T-ratio** | **P-value** | **Adjusted p-value** | **Estimate** | **T-ratio** | **P-value** | **Adjusted p-value** | **Estimate** | **T-ratio** | **P-value** | **Adjusted p-value** |
| **Pulse 1 vs. Pulse 2** | 0.01 | 0.85 | 0.34 | 1.00 | 0.04 | 1.47 | 0.14 | 0.56 | 0.02 | 1.13 | 0.26 | 1.00 | -0.05 | -1.96 | 0.050 | 0.15 |
| **Pulse 1 vs. Pulse 3** | -0.001 | -0.04 | 0.97 | 1.00 | 0.03 | 1.25 | 0.21 | 0.56 | 0.01 | 0.42 | 0.68 | 1.00 | -0.06 | -2.66 | 0.008 | **0.032** |
| **Pulse 1 vs. Pulse 4** | -0.003 | -0.21 | 0.83 | 1.00 | 0.03 | 1.36 | 0.17 | 0.56 | 0.02 | 1.38 | 0.17 | 0.84 | -0.03 | -1.19 | 0.23 | 0.46 |
| **Pulse 1 vs. Pulse 5** | 0.001 | 0.07 | 0.94 | 1.00 | 0.04 | 1.84 | 0.066 | 0.33 | 0.01 | 0.85 | 0.40 | 1.00 | -0.004 | -0.15 | 0.88 | 0.88 |
| **Pulse 1 vs. Pulse 6** | -0.01 | -0.69 | 0.49 | 1.00 | -0.001 | -0.04 | 0.97 | 0.97 | -0.01 | -0.33 | 0.74 | 1.00 | -0.07 | -3.10 | 0.002 | **0.0098** |
| **ADHD vs. autism only** | | | **Estimate** | | | **T-ratio** | | | **P-value** | | | **Adjusted p-value** | | | | |
| **Pulse 1 vs. Pulse 1** | | | -0.003 | | | -0.07 | | | 0.95 | | | 1.00 | | | | |
| **Pulse 2 vs. Pulse 2** | | | 0.02 | | | 0.39 | | | 0.70 | | | 1.00 | | | | |
| **Pulse 3 vs. Pulse 3** | | | 0.03 | | | 0.54 | | | 0.59 | | | 1.00 | | | | |
| **Pulse 4 vs. Pulse 4** | | | 0.03 | | | 0.65 | | | 0.52 | | | 1.00 | | | | |
| **Pulse 5 vs. Pulse 5** | | | 0.04 | | | 0.79 | | | 0.43 | | | 1.00 | | | | |
| **Pulse 6 vs. Pulse 6** | | | 0.01 | | | 0.12 | | | 0.90 | | | 1.00 | | | | |
| **ID vs. autism only** | | |  | | | | | | | | | | | | | |
| **Pulse 1 vs. Pulse 1** | | | 0.03 | | | 0.68 | | | 0.50 | | | 1.00 | | | | |
| **Pulse 2 vs. Pulse 2** | | | 0.04 | | | 0.86 | | | 0.39 | | | 1.00 | | | | |
| **Pulse 3 vs. Pulse 3** | | | 0.04 | | | 0.88 | | | 0.38 | | | 1.00 | | | | |
| **Pulse 4 vs. Pulse 4** | | | 0.06 | | | 1.35 | | | 0.18 | | | 1.00 | | | | |
| **Pulse 5 vs. Pulse 5** | | | 0.04 | | | 1.02 | | | 0.31 | | | 1.00 | | | | |
| **Pulse 6 vs. Pulse 6** | | | 0.03 | | | 0.79 | | | 0.43 | | | 1.00 | | | | |
| **ADHD & ID vs. autism** | | |  | | | | | | | | | | | | | |
| **Pulse 1 vs. Pulse 1** | | | 0.01 | | | 0.28 | | | 0.78 | | | 1.00 | | | | |
| **Pulse 2 vs. Pulse 2** | | | -0.05 | | | -0.92 | | | 0.36 | | | 1.00 | | | | |
| **Pulse 3 vs. Pulse 3** | | | -0.05 | | | -1.00 | | | 0.32 | | | 1.00 | | | | |
| **Pulse 4 vs. Pulse 4** | | | -0.01 | | | -0.24 | | | 0.81 | | | 1.00 | | | | |
| **Pulse 5 vs. Pulse 5** | | | 0.01 | | | 0.19 | | | 0.85 | | | 1.00 | | | | |
| **Pulse 6 vs. Pulse 6** | | | -0.05 | | | -1.02 | | | 0.31 | | | 1.00 | | | | |

**Table S8A | Short-term habituation of the acoustically evoked eyelid startle response using a rhythmic stimulus pattern in children with autism with and without co-occurring conditions.**

All statistical comparisons done using an ANOVA on Linear Mixed-Effect Model.

P-value adjustment: Bonferroni-Holm method for five tests for within group comparisons and six tests for between group comparisons.

NEC = Normalized Eyelid Closure, SD = standard deviation, ADHD = Attention Deficit Hyperactivity Disorder, ID = Intellectual Disability.

| **Random stimulus pattern** | | | | | | | | | | | | | | | | |
| --- | --- | --- | --- | --- | --- | --- | --- | --- | --- | --- | --- | --- | --- | --- | --- | --- |
|  | **Eyelid startle amplitude (mean NEC ± SD)** | | | | | | | | | | | | | | | |
| **Pulse number** | **Autism only (n = 81)** | | | | **ADHD (n = 30)** | | | | **ID (n = 56)** | | | | **ADHD & ID (n = 31)** | | | |
| **1** | 0.28 (± 0.31) | | | | 0.27 (± 0.33) | | | | 0.23 (± 0.30) | | | | 0.25 (± 0.32) | | | |
| **2** | 0.26 (± 0.34) | | | | 0.30 (± 0.37) | | | | 0.22 (± 0.32) | | | | 0.27 (± 0.36) | | | |
| **3** | 0.30 (± 0.35) | | | | 0.30 (± 0.37) | | | | 0.22 (± 0.33) | | | | 0.34 (± 0.37) | | | |
| **4** | 0.24 (± 0.33) | | | | 0.25 (± 0.35) | | | | 0.20 (± 0.32) | | | | 0.25 (± 0.36) | | | |
| **5** | 0.28 (± 0.35) | | | | 0.27 (± 0.38) | | | | 0.23 (± 0.35) | | | | 0.32 (± 0.35) | | | |
| **6** | 0.24 (± 0.33) | | | | 0.23 (± 0.27) | | | | 0.20 (± 0.33) | | | | 0.27 (± 0.35) | | | |
| **Main effect** | Pulse: F_5,9388_ = 9.69, p < 0.0001 | | | | | | | | | | | | | | | |
| **Main effect** | Co-occurring condition: F_3,193_ = 0.52, p = 0.67 | | | | | | | | | | | | | | | |
| **Interaction** | Pulse x co-occurring condition: F_15,9388_ = 1.10, p = 0.35 | | | | | | | | | | | | | | | |
|  | **Pairwise comparisons** | | | | | | | | | | | | | | | |
|  | **Estimate** | **T-ratio** | **P-value** | **Adjusted p-value** | **Estimate** | **T-ratio** | **P-value** | **Adjusted p-value** | **Estimate** | **T-ratio** | **P-value** | **Adjusted p-value** | **Estimate** | **T-ratio** | **P-value** | **Adjusted p-value** |
| **Pulse 1 vs. Pulse 2** | 0.03 | 1.80 | 0.072 | 0.22 | 0.03 | 1.13 | 0.26 | 0.91 | 0.01 | 0.37 | 0.71 | 1.00 | -0.02 | -0.75 | 0.45 | 1.00 |
| **Pulse 1 vs. Pulse 3** | -0.02 | -1.06 | 0.29 | 0.58 | -0.03 | -1.33 | 0.18 | 0.91 | 0.004 | 0.22 | 0.83 | 1.00 | -0.09 | -3.58 | 0.0003 | 0.0017 |
| **Pulse 1 vs. Pulse 4** | 0.04 | 2.70 | 0.007 | 0.028 | 0.02 | 0.83 | 0.41 | 0.91 | 0.02 | 1.39 | 0.16 | 0.66 | 0.002 | 0.09 | 0.93 | 1.00 |
| **Pulse 1 vs. Pulse 5** | 0.003 | 0.21 | 0.83 | 0.83 | -0.01 | -0.27 | 0.79 | 0.91 | -0.002 | -0.13 | 0.90 | 1.00 | -0.06 | -2.65 | 0.008 | 0.032 |
| **Pulse 1 vs. Pulse 6** | 0.05 | 3.05 | 0.002 | **0.011** | 0.03 | 1.30 | 0.19 | 0.91 | 0.03 | 1.62 | 0.11 | 0.53 | -0.02 | -0.84 | 0.40 | 1.00 |
| **ADHD vs. autism only** | | | **Estimate** | | | **T-ratio** | | | **P-value** | | | **Adjusted p-value** | | | | |
| **Pulse 1 vs. Pulse 1** | | | -0.02 | | | -0.46 | | | 0.65 | | | 1.00 | | | | |
| **Pulse 2 vs. Pulse 2** | | | -0.02 | | | -0.48 | | | 0.63 | | | 1.00 | | | | |
| **Pulse 3 vs. Pulse 3** | | | -0.01 | | | -0.12 | | | 0.91 | | | 1.00 | | | | |
| **Pulse 4 vs. Pulse 4** | | | -0.003 | | | -0.05 | | | 0.96 | | | 1.00 | | | | |
| **Pulse 5 vs. Pulse 5** | | | -0.01 | | | -0.26 | | | 0.80 | | | 1.00 | | | | |
| **Pulse 6 vs. Pulse 6** | | | -0.01 | | | -0.18 | | | 0.86 | | | 1.00 | | | | |
| **ID vs. autism only** | | |  | | | | | | | | | | | | | |
| **Pulse 1 vs. Pulse 1** | | | -0.04 | | | -1.08 | | | 0.28 | | | 1.00 | | | | |
| **Pulse 2 vs. Pulse 2** | | | -0.02 | | | -0.57 | | | 0.57 | | | 1.00 | | | | |
| **Pulse 3 vs. Pulse 3** | | | -0.06 | | | -1.57 | | | 0.12 | | | 0.71 | | | | |
| **Pulse 4 vs. Pulse 4** | | | -0.03 | | | -0.68 | | | 0.49 | | | 1.00 | | | | |
| **Pulse 5 vs. Pulse 5** | | | -0.04 | | | -0.94 | | | 0.35 | | | 1.00 | | | | |
| **Pulse 6 vs. Pulse 6** | | | -0.03 | | | -0.65 | | | 0.52 | | | 1.00 | | | | |
| **ADHD & ID vs. autism** | | |  | | | | | | | | | | | | | |
| **Pulse 1 vs. Pulse 1** | | | -0.03 | | | -0.63 | | | 0.53 | | | 1.00 | | | | |
| **Pulse 2 vs. Pulse 2** | | | 0.01 | | | 0.30 | | | 0.76 | | | 1.00 | | | | |
| **Pulse 3 vs. Pulse 3** | | | 0.04 | | | 0.81 | | | 0.42 | | | 1.00 | | | | |
| **Pulse 4 vs. Pulse 4** | | | 0.01 | | | 0.17 | | | 0.87 | | | 1.00 | | | | |
| **Pulse 5 vs. Pulse 5** | | | 0.04 | | | 0.75 | | | 0.46 | | | 1.00 | | | | |
| **Pulse 6 vs. Pulse 6** | | | 0.04 | | | 0.74 | | | 0.46 | | | 1.00 | | | | |

**Table S8B | Short-term habituation of the acoustically evoked eyelid startle response using a random stimulus pattern in autistic children with and without co-occurring conditions.**

All statistical comparisons done using an ANOVA on Linear Mixed-Effect Model.

P-value adjustment: Bonferroni-Holm method for five tests for within group comparisons and six tests for between group comparisons.

NEC = Normalized Eyelid Closure, SD = standard deviation, ADHD = Attention Deficit Hyperactivity Disorder, ID = Intellectual Disability.

|  | **Cumulative sum of eyelid amplitude (mean NEC ± SD)** | | | | **Main effect** | **B** | **SE** | **CI low** | **CI high** | **T-statistic** | **P-value** |
| --- | --- | --- | --- | --- | --- | --- | --- | --- | --- | --- | --- |
| **Stimulus pattern** | Autism only  (n = 81) | ADHD (n = 29) | ID (n = 56) | ADHD & ID  (n = 30) |  |  |  |  |  |  |  |
| **Rhythmic** | 76.59 (± 106.24) | 86.59 (± 106.52) | 70.68 (± 106.54) | 91.82 (± 118.90) | ADHD | 10.37 | 13.02 | -15.32 | 36.05 | 0.80 | 0.43 |
|  |  |  |  |  | ID | -4.94 | 10.45 | -25.55 | 15.68 | -0.47 | 0.64 |
|  |  |  |  |  | ADHD & ID | 16.02 | 12.87 | -9.37 | 41.41 | 1.24 | 0.22 |
|  | **SD of max cumulative sum** | | | |  |  |  |  |  |  |  |
|  | 84.60 (± 48.41) | 80.65 (± 34.03) | 87.57 (± 50.86) | 93.14 (± 55.75) | ADHD | -3.95 | 10.37 | -24.40 | 16.50 | -0.38 | 0.70 |
|  |  |  |  |  | ID | 2.96 | 8.43 | -13.66 | 19.59 | 0.35 | 0.73 |
|  |  |  |  |  | ADHD & ID | 8.53 | 10.25 | -11.67 | 28.74 | 0.83 | 0.41 |
| **Random** | **Cumulative sum of eyelid amplitude (mean NEC ± SD)** | | | |  |  |  |  |  |  |  |
|  | Autism only  (n = 81) | ADHD (n = 30) | ID (n = 56) | ADHD & ID  (n = 31) | ADHD | 10.92 | 12.86 | -14.45 | 36.29 | 0.85 | 0.40 |
|  | 80.65 (± 114.64) | 90.37 (± 122.78) | 68.29 (± 108.00) | 102.07 (± 115.51) | ID | -11.45 | 10.37 | -31.90 | 9.01 | -1.10 | 0.27 |
|  |  |  |  |  | ADHD & ID | 21.50 | 12.62 | -3.39 | 46.38 | 1.70 | 0.090 |
|  | **SD of max cumulative sum** | | | |  |  |  |  |  |  |  |
|  | 86.18 (± 54.01) | 100.34 (± 63.08) | 85.71 (± 47.11) | 95.92 (± 52.67) | ADHD | 14.16 | 11.42 | -8.37 | 36.68 | 1.24 | 0.22 |
|  |  |  |  |  | ID | -0.47 | 9.29 | -18.79 | 17.85 | -0.05 | 0.96 |
|  |  |  |  |  | ADHD & ID | 9.74 | 11.29 | -12.52 | 32.00 | 0.86 | 0.39 |

**Table S9 | Maximum cumulative sum of eyelid closure amplitude and the associated variability for rhythmic and random stimulus patterns in children with autism with and without co-occurring conditions.**

All statistical comparisons done using Linear Mixed-Effect Models.

NEC = Normalized Eyelid Closure, CI = 95% confidence interval, SD = Standard Deviation, ADHD = Attention Deficit Hyperactivity Disorder, ID = Intellectual Disability.

| **Behavioral response** | **Mean ± SD** | | | | **Main effect** | **B** | **SE** | **OR** | **CI low** | **CI high** | **T-statistic** | **Adjusted R^2^** | **P-value** | **Adjusted p-value** |
| --- | --- | --- | --- | --- | --- | --- | --- | --- | --- | --- | --- | --- | --- | --- |
|  | **Autism only (n = 100)** | **ADHD**  **(n = 34)** | **ID (n = 69)** | **ADHD & ID (n = 50)** |  |  |  |  |  |  |  |  |  |  |
| **Screen avoidance (% of frames)** | 11.81 (± 12.63) | 8.70 (± 10.79) | 7.58 (± 8.67) | 10.24 (± 12.66) | ADHD | -0.67 | 0.31 | 0.51 | 0.28 | 0.94 | -2.17 | 0.013 | 0.030 | 0.27 |
|  |  |  |  |  | ID | -0.50 | 0.23 | 0.61 | 0.38 | 0.96 | -2.15 | 0.013 | 0.031 | 0.31 |
|  |  |  |  |  | ADHD & ID | -0.15 | 0.26 | 0.86 | 0.52 | 1.44 | -0.57 | 0.013 | 0.57 | 1.00 |
| **Headphone touches (% of trials)** | 12.08 (± 18.31) | 10.15 (± 14.11) | 11.17 (± 16.66) | 9.65 (± 14.87) | ADHD | -0.31 | 0.39 | 0.73 | 0.34 | 1.58 | -0.79 | 0.001 | 0.43 | 1.00 |
|  |  |  |  |  | ID | -0.05 | 0.31 | 0.95 | 0.52 | 1.73 | -0.16 | 0.001 | 0.87 | 1.00 |
|  |  |  |  |  | ADHD & ID | -0.06 | 0.35 | 0.94 | 0.48 | 1.85 | -0.18 | 0.001 | 0.86 | 1.00 |
| **Non-syllabic vocalizations (% of trials)** | 10.93 (± 14.77) | 9.42 (± 14.31) | 9.56 (± 14.27) | 15.93 (± 18.39) | ADHD | -0.03 | 0.41 | 0.97 | 0.44 | 2.15 | -0.08 | 0.019 | 0.93 | 1.00 |
|  |  |  |  |  | ID | -0.44 | 0.33 | 0.65 | 0.34 | 1.23 | -1.34 | 0.019 | 0.18 | 1.00 |
|  |  |  |  |  | ADHD & ID | 0.65 | 0.35 | 1.92 | 0.96 | 3.85 | 1.85 | 0.019 | 0.064 | 0.51 |
|  |  |  |  |  |  |  |  | **β** |  |  |  |  |  |  |
| **AP postural stability (mean SD over all trials)** | 4.24 (± 2.86) | 4.36 (± 2.83) | 4.51 (± 3.05) | 4.00 (± 2.45) | ADHD | 0.08 | 0.16 | 0.10 | -0.23 | 0.39 | 0.51 | -0.008 | 0.61 | 1.00 |
|  |  |  |  |  | ID | 0.11 | 0.12 | 0.13 | -0.14 | 0.35 | 0.85 | -0.008 | 0.34 | 1.00 |
|  |  |  |  |  | ADHD & ID | 0.01 | 0.14 | 0.01 | -0.26 | 0.28 | 0.06 | -0.008 | 0.95 | 1.00 |
| **Head rotations (% of trials)** | 36.23 (± 24.20) | 37.86 (± 21.36) | 41.57 (± 25.94) | 45.04 (± 28.07) | ADHD | 0.14 | 0.17 | 0.16 | -0.19 | 0.47 | 0.83 | 0.016 | 0.41 | 1.00 |
|  |  |  |  |  | ID | 0.26 | 0.13 | 0.30 | 0.00 | 0.52 | 1.96 | 0.016 | 0.051 | 0.46 |
|  |  |  |  |  | ADHD & ID | 0.35 | 0.15 | 0.41 | 0.06 | 0.64 | 2.36 | 0.016 | 0.019 | 0.17 |
| **Mouth openings (mean area over all trials)** | 8.40 (± 5.60) | 10.90 (± 7.69) | 8.60 (± 6.90) | 12.66 (± 8.13) | ADHD | 0.32 | 0.18 | 0.35 | -0.03 | 0.67 | 1.82 | 0.048 | 0.070 | 0.56 |
|  |  |  |  |  | ID | 0.00 | 0.14 | -0.004 | -0.28 | 0.27 | -0.03 | 0.048 | 0.98 | 1.00 |
|  |  |  |  |  | ADHD & ID | 0.54 | 0.16 | 0.59 | 0.24 | 0.85 | 3.50 | 0.048 | 0.001 | **0.0056** |
| **Pupil range (mean horizontal movement over all trials)** | 13.87 (± 4.21) | 13.27 (± 3.59) | 14.84 (± 4.26) | 14.71 (± 3.94) | ADHD | -0.60 | 0.81 | -0.15 | -2.20 | 1.00 | -0.74 | 0.007 | 0.46 | 1.00 |
|  |  |  |  |  | ID | 0.97 | 0.64 | 0.24 | -0.29 | 2.23 | 1.51 | 0.007 | 0.13 | 0.92 |
|  |  |  |  |  | ADHD & ID | 0.84 | 0.71 | 0.20 | -0.56 | 2.24 | 1.18 | 0.007 | 0.24 | 1.00 |
| **Side eye** | 0.13 (± 0.09) | 0.15 (± 0.11) | 0.13 (± 0.08) | 0.13 (± 0.06) | ADHD | 0.18 | 0.18 | 0.20 | -0.18 | 0.54 | 1.00 | -0.006 | 0.32 | 1.00 |
|  |  |  |  |  | ID | -0.03 | 0.14 | -0.04 | -0.32 | 0.25 | -0.24 | -0.006 | 0.81 | 1.00 |
|  |  |  |  |  | ADHD & ID | -0.02 | 0.16 | -0.02 | -0.33 | 0.29 | -0.14 | -0.006 | 0.890 | 1.00 |
| **Baseline regression intercept** | 7.22 (± 4.40) | 10.36 (± 6.60) | 8.86 (± 7.27) | 8.48 (± 4.54) | ADHD | 0.55 | 0.20 | 0.53 | 0.15 | 0.95 | 2.71 | 0.020 | 0.007 | 0.073 |
|  |  |  |  |  | ID | 0.25 | 0.16 | 0.25 | -0.06 | 0.57 | 1.59 | 0.020 | 0.11 | 0.91 |
|  |  |  |  |  | ADHD & ID | 0.25 | 0.18 | 0.25 | -0.09 | 0.60 | 1.44 | 0.020 | 0.15 | 0.91 |
| **Baseline regression SD** | 4.82 (± 2.25) | 5.44 (± 3.20) | 5.38 (± 2.98) | 5.54 (± 2.08) | ADHD | 0.18 | 0.19 | 0.19 | -0.20 | 0.55 | 0.94 | 0.003 | 0.35 | 1.00 |
|  |  |  |  |  | ID | 0.18 | 0.15 | 0.19 | -0.11 | 0.48 | 1.21 | 0.003 | 0.23 | 1.00 |
|  |  |  |  |  | ADHD & ID | 0.30 | 0.17 | 0.31 | -0.03 | 0.63 | 1.81 | 0.003 | 0.071 | 0.51 |

**Table S10 | Smartphone-based assessments of behavioral responses in autistic children with and without co-occurring conditions.**

Multilevel binomial logistic regression for screen avoidance, headphone touches, non-syllabic vocalizations.

Linear regression for: AP postural stability (log transformed); head rotations (log transformed); mouth openings (square root transformed); side eye (square root transformed); baseline regression intercept (square root transformed); baseline regression SD (square root transformed).

P-value adjustments: Bonferroni-Holm method for ten tests.

SD = Standard Deviation, CI = 95% confidence interval, SE = Standard Error, OR = Odds Ratio, AP = Anteroposterior.
