## supplementary methods for "Neurobehavioral Assessment of Sensorimotor Function in Autism Using Smartphone Technology"

**Experimental Setup:** During the experiment, children watched an audio-normalized movie while the app delivered auditory stimuli. The iPhone models used in this study were iPhone 12 and iPhone 13 Pro Max. For delivery of auditory cues, we used wired headphones. Children were supervised by an experimenter on-site. The test environment was monitored by the app as previously described (Boele et al., 2023). For an illustration of the experimental setup and the child being administered the test, refer to **supplementary** **figure 2**. The parents of the children pictured provided informed consent for publication of these images in an online open-access publication. For each trial, computer vision algorithms were used to track and record the position of the participant’s facial landmarks over time and to determine amplitude and timing of mouth opening, head and postural movements, as well as the eyelid closure (**figure 1A**). Eyelid position signals were captured and calculated in real time as previously described (Boele et al., 2023).

Participants were restricted to choosing the following movies: Masha et Michka, Baby Bus Episode 1, Toyor Songs, Paw Patrol, Friend City, Spirit, Pyjamasque, Baby Bus Episode 2, Super Mario, Funkie Veggies, Rainbow Caterpillar.

**Prepulse Inhibition (PPI) paradigm:** This paradigm was delivered during Test I. We studied PPI of the acoustically evoked eyelid startle response, using a 50 ms white noise audio burst at 105 dB as the pulse, accompanied by prepulse bursts categorized as weak (5% of pulse amplitude), medium (10% of pulse amplitude), and high (25% of pulse amplitude) at intensities of 65 dB, 75 dB, and 83 dB respectively. Participants completed one session which contained 34 trials. First, a total of two habituation trials containing white noise bursts of various soft intensities were presented. This allowed for the child to relax and settle into the movie. After these two trials, eight blocks of four trials were presented. Each block consisted of a pulse only trial and three prepulse-pulse trials. A prepulse always preceded the pulse by 120 ms. Intertrial interval (ITI) was set at random between 10 and 25 seconds. A training session lasted for about 15 minutes.

**Short-term habituation (short-term HAB) paradigms:** This paradigm was delivered during Test II. Children completed one session which contained 22 stimulus trials. Trials included two practice trials with weak audio pulses which allowed children to get acclimated to the stimuli. The remaining 20 trials were short-term HAB trials. For these trials, we used pulse trains with six 50 ms white noise audio pulses, at 105 dB, with the last pulse presented at 4.5 seconds. Two distinct stimulus trains each presented 10 times, in a randomized, but fixed order were used: 1) 10 rhythmic trains were delivered where the pulses were presented at a 1.33 Hz frequency. Pulse number six was delayed by a full cycle to create a missing pulse sensation (**supplementary figure 1**). 2) A more ‘random’ feeling pulse train was delivered 10 times, here the basic rhythm was still 1.33 Hz, but the pulse two to pulse three and pulse four to pulse five interval was shifted to 1.25s and 1s respectively. The ITI was set at random between 20 and 40 seconds. A training session lasted around 15 minutes.

**Analysis of neurometric signals (eyelids):** Eyeblink traces were collected using the iPhone app, as previously described (Boele et al., 2023). These raw traces were filtered using a median filter (window size = 3) applied symmetrically to preserve the temporal profile of individual blinks. To improve synchronization between the eyeblink data and stimulus timing, the filtered traces were resampled from 60 Hz to 100 Hz. This processing was applied separately to data from the left and right eyes. The resampled signals were then averaged to yield a single combined trace for each trial. To enable inter-subject comparisons, we normalized these combined traces. The normalization procedure first determined two reference points: (1) the maximum blink amplitude across all combined traces (upper limit), and (2) the median baseline value across all combined traces in the window from 1000 ms to 250 ms prior to stimulus onset (lower limit). Each trace was normalized by subtracting the lower limit and dividing by the difference between the upper and lower limits. This resulted in a normalized scale where 0 corresponds to eyes open and 1 corresponds to eyes closed. All filtering and resampling procedures were implemented using the SciPy library. All subsequent analyses were performed on this normalized data. Utilizing a combination of the smartphone’s forward-facing camera and microphone enabled us to gather precise positions regarding eyelid movement. Trials were excluded under several conditions, including pre-stimulus blinks (defined as detected peaks, using the findpeaks function from SciPy, occurring 150ms before and 30ms after the onset of the first stimulus), the absence of headphones on the child, the child’s face being out of the camera frame resulting in more than 25% missing data points in a trial or more than 10% missing data points in a trial window ranging from 150ms before to 750ms after a pulse, incorrect subject tracking, or issues with the stimuli not being delivered.

**Acoustic startle response (ASR) analysis:** This is reflexive response, for example an eyeblink, to a startling auditory stimulus. In all participants, we studied the median maximum amplitude of the eyelid startle response to a single 50 ms white noise pulse at 105 dB in an ASR window. The ASR window was determined as follows: blinks occurring 0-250ms after the pulse were detected; if fewer than 3 blinks occurred in this window, all trials (instead of just the pulse trials) were used to detect stimulus-evoked blinks. Using these detected blinks, the lower bound of the ASR window was then calculated as: 25th percentile - 1.5*IQR and the upper bound as: 75th percentile + 1.5*IQR. If the ASR window size was smaller than 100ms or larger than 200ms, the blink median ± 50ms was used to create a 100ms ASR window around the median peak time. For all other eyelid signal analyses, the same principle was used to determine the ASR window from which the maximum amplitude of NEC was calculated.

**Long-term habituation (long-term HAB) analysis:** This measure was used to describe the change in the amplitude of the ASR over the course of a PPI experiment. Pulse-only trials were paired in the order in which they occurred in the PPI experiment. We then calculated the average maximum NEC startle amplitude for each pair to investigate the startle amplitude over the full length of a test. We realize that this type of paradigm is commonly referred to as habituation, however, we refer to this as long-term HAB to avoid confusion with the short-term HAB paradigms described below. Again, response detection windows were the ASR windows described above.

**PPI analysis:** PPI is the reduction in the ASR amplitude to a normally startling stimulus in the presence of a preceding weaker stimulus. The ASR window was used as the response detection window for eyelid responses to the auditory stimuli. To analyze amplitude reduction, we used the maximum amplitude of the NEC in the ASR window and then calculated the median per trial type (pulse; pulse + prepulse 5%; pulse + prepulse 10%; pulse + prepulse 25%). We also analyzed the variability in startle responses to the PPI stimuli by calculating the standard deviation of the amplitude of NEC per trial type.

For figures depicting the PPI percentage, the following formula was used: % PPI = 100 - (mean ASR of trial type/mean ASR pulse trials x 100). To avoid extreme values, participants with a median startle amplitude (< 0.075 NEC) for pulse-only trials were excluded from figures showing percentage of PPI.

**Short-term startle habituation (short-term HAB) analysis:** We defined short-term HAB as the change in the ASR amplitude across a series of white noise pulses presented in a rhythmic or random pattern. Response detection windows were calculated like the ASR windows defined above, however, all stimulus times in all trials were used for blink detection in the short-term HAB experiment. Short-term HAB was calculated by comparing the maximum NEC startle amplitude across response windows for every pulse for both the rhythmic and random paradigm.

**Anticipatory eye blinks (AEB):** AEBs were defined as blinks in windows where no stimulus was presented but, based on the rhythmic pattern of stimulus delivery, a stimulus was expected. Interstimulus intervals (ISIs) were calculated for the first four pulse stimuli in the rhythmic short-term HAB paradigm. “Missing stimuli” were then defined as instances where no pulse noise occurred at stimulus_n-1_ + ISI. AEB windows were defined as -100ms and + 200ms around the expected (but omitted) stimulus time. The maximum NEC amplitude was then determined in these AEB windows. These windows measure the level in which participants were trying to predict the stimuli in a rhythmic fashion. (**Supplementary figure 1**)

**Cumulative sum of eyelid trace:** Eyelid traces for the short-term HAB paradigms were first baseline corrected to minimize the effect of negative values. Baselines were defined as a 1000ms window before the onset of the first pulse. The 10th percentile of this window was calculated and subtracted from the eyelid trace creating a baseline corrected trace. The cumulative sum for each short-term HAB paradigm was then calculated from this baseline corrected trace with the NumPy cumsum function. The mean of the last 10 values of the cumulative sum per short-term HAB paradigm was determined for each subject and then compared between groups. This was used as a measure to quantify both potential short-term HAB as well as anticipation. The standard deviation (SD) of this maximum cumulative sum value was also calculated for each subject and compared between groups.

**Analysis of behavioral data:** During the presentation of the stimuli, the phone’s forward-facing camera and microphone captured the child’s behavioral responses. The generated videos provided a detailed record of the participants’ facial expressions, head position and vocalizations in response to the stimuli. These videos were analyzed for neurobehavioral responses. The following six behavioristic parameters were quantified:

1. Screen avoidance: behavior was quantified as the average percentage of frames per trial where the participant’s face was not detected using the Apple Vision framework.

2. Headphone touches: trained coders observed trial videos to record instances where the child used their hands to touch any part of the headphones that were on their head.

3. Vocalizations: trained coders observed trial videos to record timing of vocalizations, as well as their categories. Vocalizations were categorized based on the nature of the sounds rather than their linguistic content (Tenenbaum et al., 2020). Non-syllabic vocalizations included nonverbal vocal sounds not forming part of words.

4. Anteroposterior postural stability: we quantified the variability in face distance from the phone's camera by calculating the size of the bounding box relative to the full field of view. The relative bounding box size was determined by normalizing the area of the bounding box around the child’s face in each video frame against the total screen area. The dimensions of the bounding box were derived from the differences in bounding coordinates, multiplied by the respective video dimensions. This proportion, rounded to three decimal places, served as a measure of the child's movement extent.

5. Head rotations: This is calculated per video frame. Each frame contains data points that form a line from the detected top of the nose down to the detected subject's chin, namely the “median line”. The roll of the head is corrected per frame. For this the two data-points close to the temples are used. The midpoint and angle of the line connecting these two points is computed and the median line landmark coordinates are rotated to correct for this angle. Then, from the corrected median line landmarks the chin landmark is used to determine its relative horizontal position between the corrected temple positions, using a range of -0.5, to +0.5. For each trial the standard deviation is calculated and averaged over the entire result. The value is then multiplied by 1000 for readability purposes. The value can be interpreted as the variation in a subject’s horizontal head rotations.

6. Mouth openings: assessing whether the child's mouth was open, was calculated by analyzing the area of the inner lips in each video frame. The process involves collecting mouth landmark data from the video frames and computing the area enclosed by the inner lip landmarks using Gauss’s area formula where the vertices are data points from the inner lips. This results in a mouth opening trace per trial which is averaged to get a mouth opening value per result and multiplied by 100 for readability purposes.

7. Pupil range: This indicates the range of horizontal pupil movements. A mean histogram for each trial type is calculated separately for the left and right pupil. The range from these histograms is determined to give a mean pupil movement range per trial type. Data from the left and right pupil is then combined and averaged over trial types to give the pupil range value which shows how much a subject moves their pupils relative to their eyes.

8. Side eye: The mean pupil movement (left pupil data and right pupil data with the sign reversed) and head rotation data are used. When the direction of head rotation and pupil movement is the same the side-eye value is 0. When the directions are opposite, the side-eye is calculated as: |head rotation| + |mean pupil movement|. Thus, a value of 1 corresponds to completely opposite head and pupil positions.

9. Baseline regression intercept and SD: The variation of the mean baseline NEC amplitude is determined over time by using a rolling window approach (window-size = 5 trials) and calculating the Median Absolute Deviation (MAD). The linear regression is calculated from the rolling MAD regression data, resulting in the intercept. Additionally, the variance from the rolling MAD regression data is calculated to determine the SD. Both the intercept and SD values are multiplied by 100 for readability purposes.

**Statistical analysis of group differences:** Statistical analysis and data visualizations were done in Python 3.11 as well as R 4.3.1. For all PPI and HAB eyelid signal analyses, trials were only considered if there were a minimum of three trials per trial type (pulse only, prepulse 5% + pulse, prepulse 10% + pulse, prepulse 25% + pulse, rhythmic short-term HAB, random short-term HAB).

For analyses with nested data, we used multilevel linear mixed-effects (LME) models because they are more robust to violations of normality assumptions, which is often the case in biological data samples. LME models can better accommodate the nested structure of our data (i.e., trial nested within session, session nested within subject, subject nested within group) and prevent data loss by using summary measures. As an added benefit, LME models are better at handling missing data points than repeated measures analysis of variance (ANOVA) models and do not require homoscedasticity as an inherent assumption (Aarts et al., 2014; Schielzeth et al., 2020). For PPI, AEB and cumulative sum LME models, subject was used as a random effect and diagnosis (or co-occurring condition; and age or sex for models investigating confounding effects) was used as a fixed effect. For all ANOVA on LME models subject was used as a random effect and fixed effects were: label, for within group PPI models; trial number*diagnosis, for long-term HAB models (or co-occurring condition; and age or sex for models investigating confounding effects); pulse number*diagnosis (or co-occurring condition; and age or sex for models investigating confounding effects). For multilevel binomial logistic regression models, a random slope was used for the effect of session (PPI or HAB) across subjects and diagnosis (or co-occurring condition; and age or sex for models investigating confounding effects) was used as a fixed effect.

The distribution of data was visually inspected using histograms. Due to clear deviations from normality the following variables were transformed based on what would allow optimal model fit determined by the output from R’s bestNormalize package: postural stability (log-transformed), head rotations (log-transformed), mouth opening (square-root transformation), side-eye (square-root transformation), baseline regression intercept and SD (square-root transformation).

Goodness-of-fit model comparison was determined by evaluating log likelihood ratio, BIC, and AIC values. The distribution of residuals was inspected visually by plotting the quantiles of standard normal versus standardized residuals (i.e. Q-Q plots). Data were considered as statistically significant if the p-value was less than 0.05. For multiple comparisons, p-values were Bonferroni-Holm adjusted for the number of comparisons made.
