## supplementary figures for "Neurobehavioral Assessment of Sensorimotor Function in Autism Using Smartphone Technology"

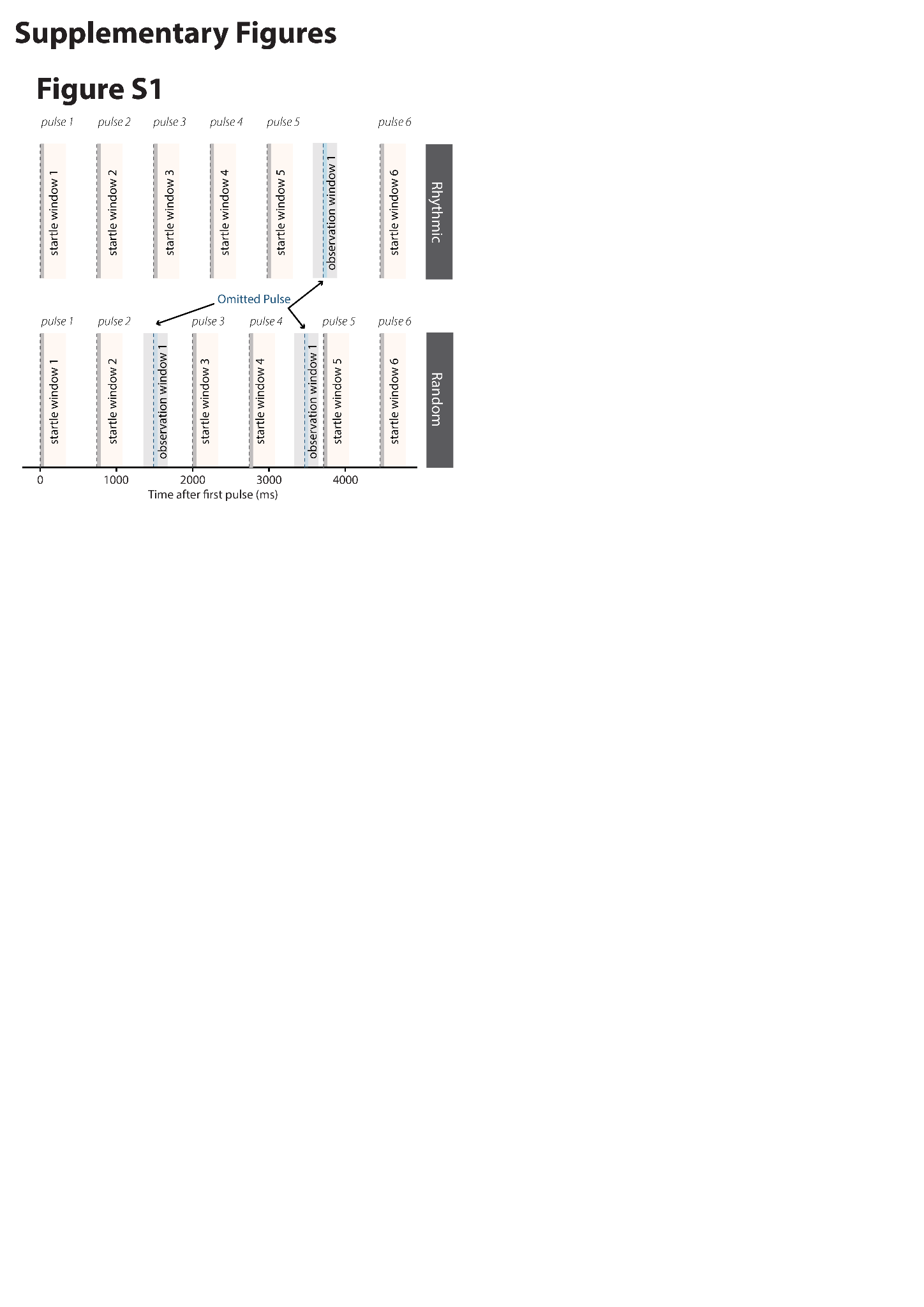

**Figure S1 | Short-term HAB experimental design.** Two stimulus patterns were delivered namely a rhythmic and random pattern. For each, six white noise pulses of 50ms each, onset indicated by dotted grey line, duration indicated by light grey block, were delivered. Startle amplitudes in startle window 1 were compared to startle amplitudes in the subsequent five windows. Startle window duration is indicated in yellow. Observation windows indicated in grey were windows where a pulse was expected (dotted blue line) but not presented.

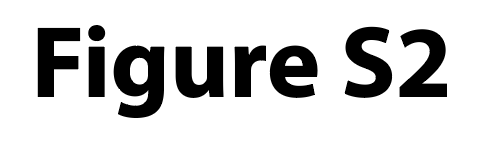

For access to this image, please contact the corresponding authors

**Figure S2 | Conducting BlinkLab experiments (A, C)** Experimenter readies the app for a child, ensuring a quiet environment with suitable lighting and correct headphone positioning. The child is comfortably seated, ready for the test. **(B, D)** The child watches an age-appropriate movie on a smartphone with auditory stimuli presented via headphones. During each trial, facial landmark detection tracks and records the child’s facial features, allowing for the assessment of eyelid closure amplitude and timing as well as postural, head, facial, and mouth movements. The researcher and parents of the children pictured provided informed consent for publication of these images in an online open-access publication.

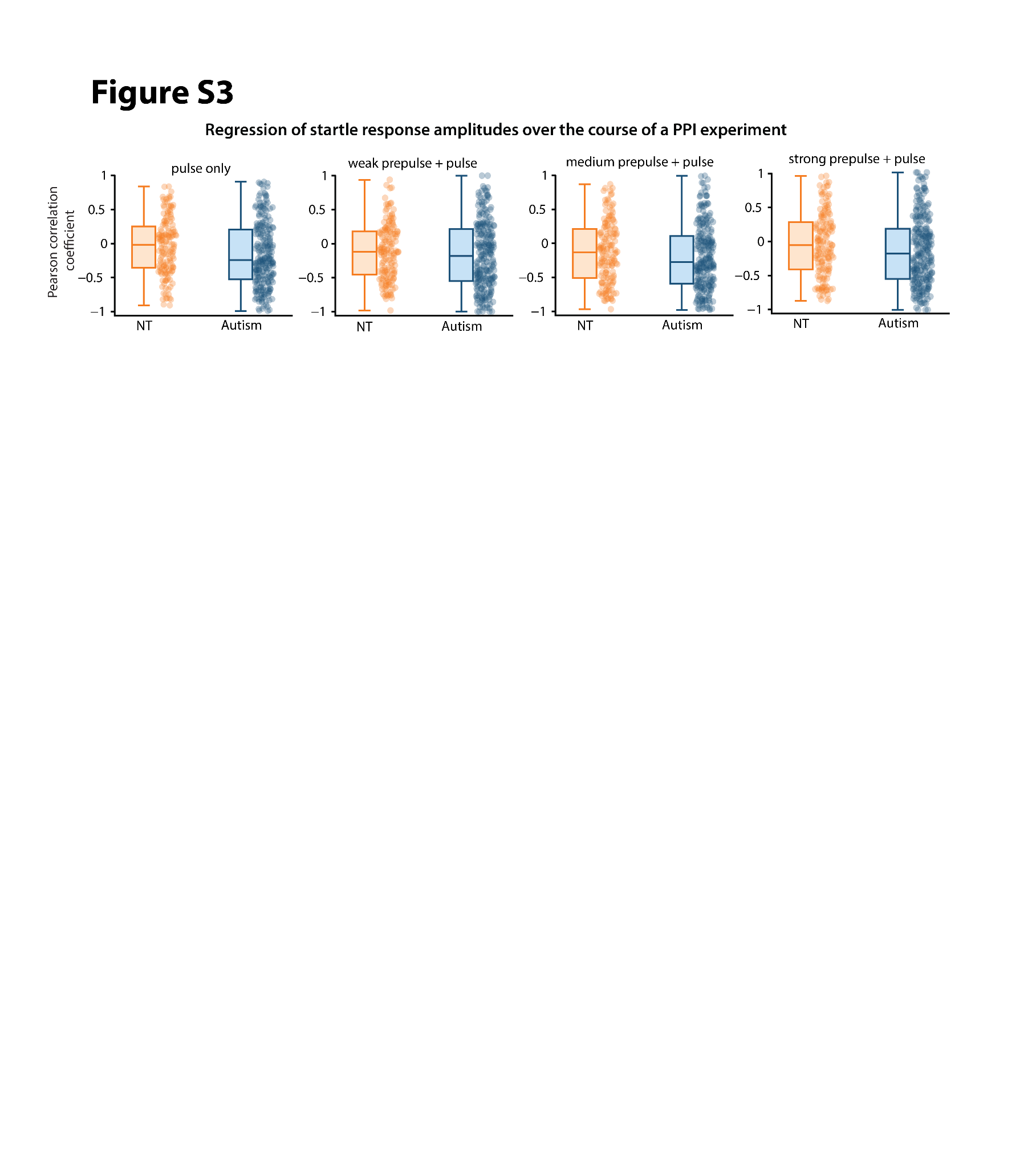

**Figure S3 | Regression of smartphone-mediated startle response amplitudes over the course of a prepulse inhibition experiment.** Group-averaged Pearson correlation coefficients from regression of eyelid closure amplitudes over the course of a PPI experiment for children with autism and NT children by trial-type. Pearson correlation coefficients differed significantly between autistic and NT children for pulse-only trials

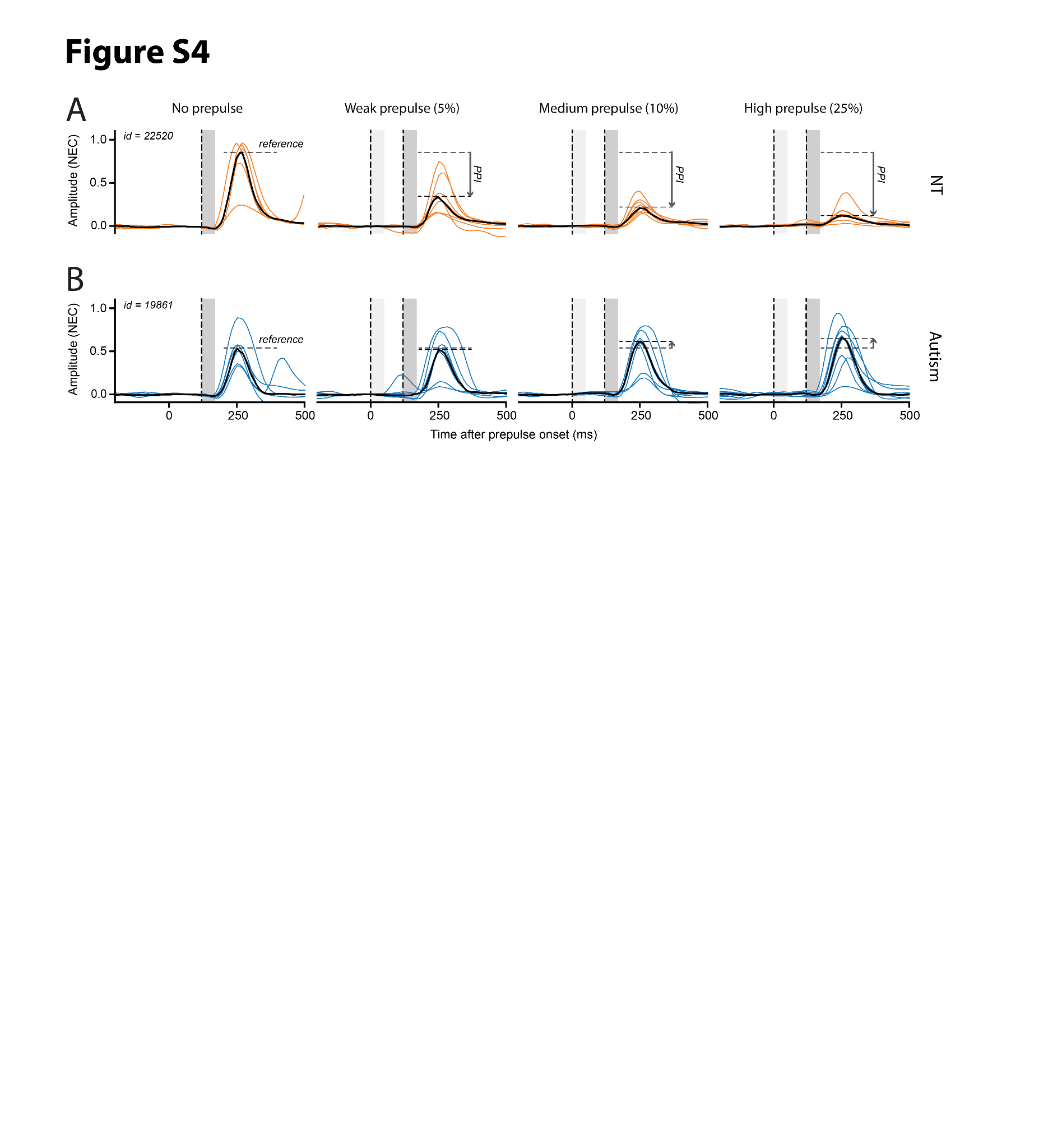

**Figure S4 | Smartphone-mediated prepulse inhibition of the acoustically evoked eyelid startle response in a neurotypical child and a child with ASD.** Response amplitude to auditory stimuli is expressed as normalized eyelid closure (NEC) on a scale from 0 (fully open) to 1 (fully closed) at a participant level. The thin colored traces correspond to individual trials within a single session; the black trace is the mean of all these trials. Left vertical dash line indicates the onset of the auditory prepulse, the right one indicates the onset of the pulse. Dark gray shaded vertical bar indicates the duration of the pulse, the light gray one indicates the duration of the prepulse (both 50 ms). The stimulus was a 50 ms white noise pulse. The prepulse was a softer white noise sound presented at an intensity of 5, 10, or 25% of pulse intensity, which remains constant at 100%. The prepulse onset always precedes the pulse onset by 120 ms. The reference vertical dashed line is the maximum amplitude of the mean pulse-only trace. The ID is referring to the participant’s id in our study. **(A)** Data from a neurotypical (NT) child showing a reduction in mean eyelid startle amplitude, i.e. prepulse inhibition, with increasing intensities of the prepulse. **(B)** A child with autism showing no reduction and even a slight increase in mean startle amplitude with increasing intensities of the prepulse.

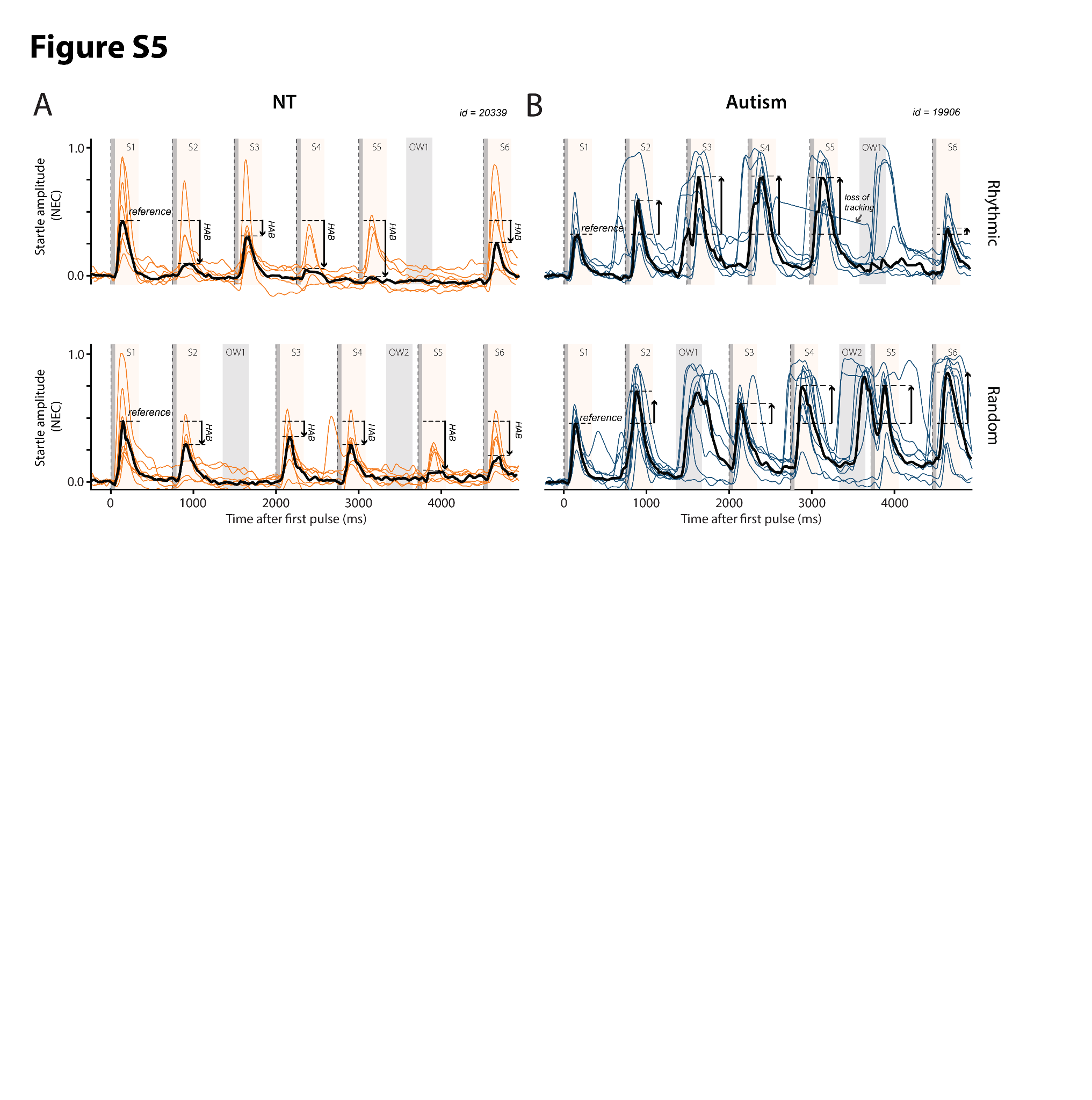

**Figure S5 | Smartphone-mediated short-term startle habituation of the acoustically evoked eyelid startle response in rhythmic and random stimulus patterns for a neurotypical child and a child with autism.** Response amplitude to auditory stimuli expressed as normalized eyelid closure (NEC) on a scale from 0 (fully open) to 1 (fully closed) at the participant level. The colored traces correspond to individual trials within a single session, while the black traces are the mean of all these trials. The onset of the auditory stimulus is the dashed line. The gray shaded vertical bar represents the duration of the stimulus at 50 ms. The rhythmic pulse train consisted of 6 pulses delivered over 4.5s, which is 750 ms between each pulse, pulse number six was delayed by 750 ms. The random pulse train still consisted of six pulses delivered over 4.5s, but pulse two to pulse three and pulse four to pulse five intervals were shifted to 1.25s and 1s respectively. The ID is referring to the participant’s id in our study. (**A**) Data from a neurotypical (NT) child showing weak short-term habituation over the six consecutive trials in both the random and the rhythmic protocol. (**B**) A child with autism showing no short-term habituation and a strong anticipation.
